# Cross-ancestry, cell-type-informed atlas of gene, isoform, and splicing regulation in the developing human brain

**DOI:** 10.1101/2023.03.03.23286706

**Authors:** Cindy Wen, Michael Margolis, Rujia Dai, Pan Zhang, Pawel F. Przytycki, Daniel D. Vo, Arjun Bhattacharya, Nana Matoba, Chuan Jiao, Minsoo Kim, Ellen Tsai, Celine Hoh, Nil Aygün, Rebecca L. Walker, Christos Chatzinakos, Declan Clarke, Henry Pratt, PsychENCODE Consortium, Mette A. Peters, Mark Gerstein, Nikolaos P. Daskalakis, Zhiping Weng, Andrew E. Jaffe, Joel E. Kleinman, Thomas M. Hyde, Daniel R. Weinberger, Nicholas J. Bray, Nenad Sestan, Daniel H. Geschwind, Kathryn Roeder, Alexander Gusev, Bogdan Pasaniuc, Jason L. Stein, Michael I. Love, Katherine S. Pollard, Chunyu Liu, Michael J. Gandal

## Abstract

Genomic regulatory elements active in the developing human brain are notably enriched in genetic risk for neuropsychiatric disorders, including autism spectrum disorder (ASD), schizophrenia, and bipolar disorder. However, prioritizing the specific risk genes and candidate molecular mechanisms underlying these genetic enrichments has been hindered by the lack of a single unified large-scale gene regulatory atlas of human brain development. Here, we uniformly process and systematically characterize gene, isoform, and splicing quantitative trait loci (xQTLs) in 672 fetal brain samples from unique subjects across multiple ancestral populations. We identify 15,752 genes harboring a significant xQTL and map 3,739 eQTLs to a specific cellular context. We observe a striking drop in gene expression and splicing heritability as the human brain develops. Isoform-level regulation, particularly in the second trimester, mediates the greatest proportion of heritability across multiple psychiatric GWAS, compared with eQTLs. Via colocalization and TWAS, we prioritize biological mechanisms for ∼60% of GWAS loci across five neuropsychiatric disorders, nearly two-fold that observed in the adult brain. Finally, we build a comprehensive set of developmentally regulated gene and isoform co-expression networks capturing unique genetic enrichments across disorders. Together, this work provides a comprehensive view of genetic regulation across human brain development as well as the stage- and cell type-informed mechanistic underpinnings of neuropsychiatric disorders.

**Graphical Abstract:** 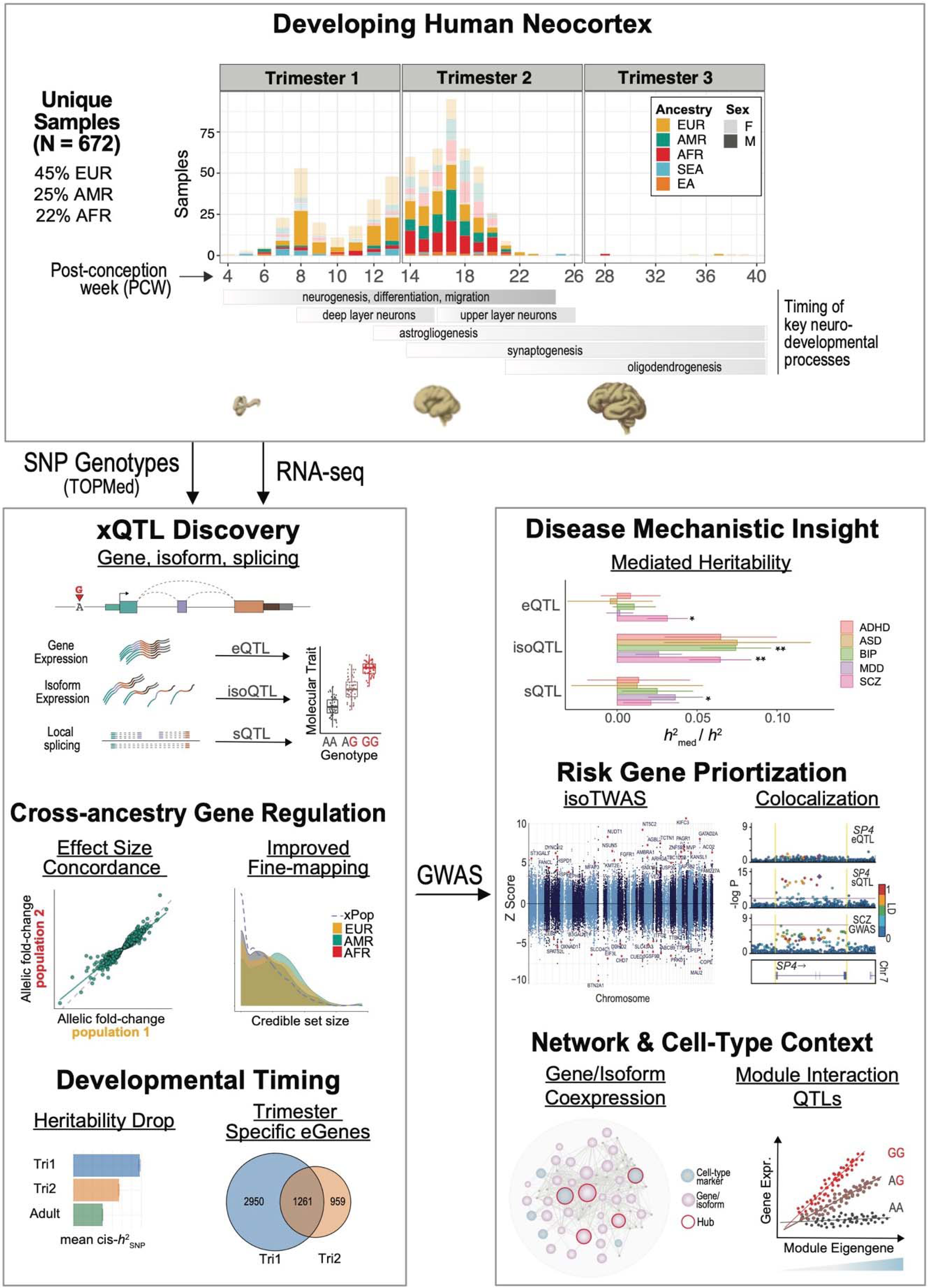

## Main Text

Thousands of genetic risk loci have now been robustly associated with neurodevelopmental and psychiatric disorders by large-scale GWAS (*1, 2*). However, as most associated GWAS variants reside within non-coding regions of the human genome, often in large linkage disequilibrium (LD) blocks, the true underlying causal variant(s) and their target gene(s) remain largely unknown. Consequently, the critical defining obstacle of the post-GWAS era is to pinpoint the specific, locus-level molecular impact of GWAS variants at scale (*3*). As risk loci are enriched in known regulatory regions of the human genome (*4–6*), one major approach to address this challenge has been to connect risk variants with tissue-specific reference panels of expression quantitative trait loci (eQTLs), through statistical colocalization, transcriptome-wide association (TWAS), and related approaches (*7–9*). This has prompted several large-scale efforts (*10–16*) to generate comprehensive functional genomic compendia connecting population-level allelic variation with gene expression profiles in the human brain. Yet, while these resources have provided biologically interpretable and meaningful annotations for dozens of neuropsychiatric risk loci, the majority remain mechanistically unannotated (*15, 17*).

Gene regulation is known to be highly dependent on the specific underlying developmental stage, tissue, and cellular context (*7, 17–20*), and increasing evidence implicates the developing human brain in the genetic risk for neuropsychiatric disorders, including autism spectrum disorder (ASD) and schizophrenia (SCZ) (*21–23*). Consequently, several efforts have begun to characterize the genetic control of gene expression during human brain development, finding that gene regulation is tightly controlled during brain development and enriched for neuropsychiatric risk (*11, 12, 24–28*). However, as these studies are individually small in sample size, the power to pinpoint the developmental timing or elucidate the full extent of gene regulation in the developing human brain has been limited.

The regulation of transcript-isoform structure and diversity, through alternative local splicing and differences in transcriptional start and termination sites, is also known to be a mechanism critical for human brain development that is implicated in disease pathogenesis (*14, 29–32*). Indeed, relative to other tissues and species, the landscape of alternative splicing is particularly extensive in the human brain, and its regulation is notably distinct from that of gene expression (*16, 33*). While individual studies have begun to investigate splicing and isoform regulation (e.g., sQTL and isoQTLs) in the developing brain (*24, 25*), a lack of uniform data processing has precluded a systematic characterization of these critical mechanisms and their potential relationships with genetic risk factors for neuropsychiatric disorders.

To address these critical gaps, here we present the most comprehensive investigation into the genetic regulation of the transcriptome during human brain development by uniformly processing data from 672 distinct samples spanning 4-39 post-conception weeks (PCW), most of which are within the first and second trimesters. The full integrated dataset is highly diverse, comprising several major continental ancestries including European (EUR; 45%), Hispanic/Latino (AMR; 25%), African (AFR; 22%), and East Asian/Southeast Asian (EA/SEA; 8%). This well-powered, cross-population resource provides an unprecedented view of the developmental timing and regulatory landscape of human brain development at gene, isoform, and splicing levels. We observe a substantial drop in the heritability of gene expression and local splicing with development, particularly from 10-18 PCW. Yet, psychiatric GWAS signals appear more concentrated within the second trimester. We find that isoform-level regulation mediates substantially more psychiatric GWAS heritability than gene or local splicing regulation. We are able to connect ∼60% of GWAS loci of five neuropsychiatric disorders with specific molecular mechanisms through colocalization and TWAS. Finally, we contextualize these risk mechanisms within a comprehensive set of gene and isoform-level co-expression networks across fetal brain development.

### Large-Scale Functional Genomic Interrogation of Human Brain Development

Here, we present a comprehensive investigation of gene expression, splicing, and isoform regulation during human brain development by integrating data across five individual cohorts (*12, 24–27*) (**Fig. S1**) comprising 672 distinct donors. Following uniform data processing, including imputation into the multi-ancestry TOPMed reference panel (*34*), and strict quality control (**Fig. S2**; Methods), we retained 654 samples with matched genotype and cortical RNA-seq data spanning 4-39 PCW, the majority falling within the first two trimesters (Tri1, n=216; Tri2, n=433; **Table S1**). Altogether, our large sample size enabled detection of 10,094 genes with at least one *cis*-eQTL at FDR < 0.05 (termed “eGenes”), following permutation-based and FDR correction for multiple comparisons while accounting for local LD structure (**Fig. 1A**; **Fig. S3**; **Table S1**; Methods).

**Fig. 1.**
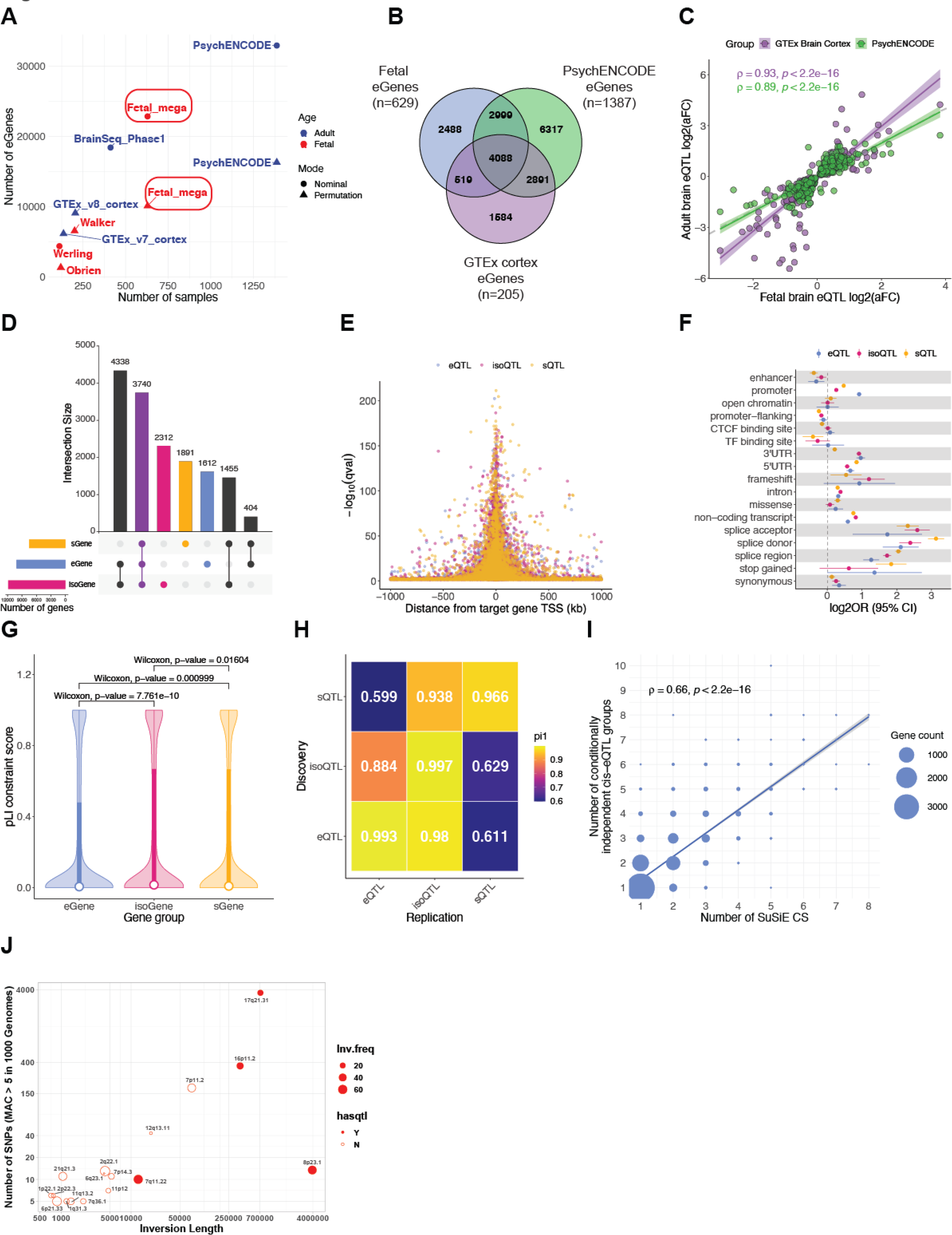
The landscape of gene, splicing, and isoform regulation in the developing human brain. **(A)** The number of eGenes discovered here versus sample size, compared with other human brain studies. **(B)** Overlap of eGenes between fetal brain (n=629), GTEx v8 Brain Cortex (n=205), and PsychENCODE (n=1,387). **(C)** Correlation of eQTL effect size, measured by allelic fold change (aFC), between fetal and adult brain. Each dot is a shared pair of eGene-primary eQTL between fetal brain and GTEx (247 pairs) or PsychENCODE (253 pairs). **(D)** Overlap among eGenes, isoGenes, and sGenes. **(E)** Distance between the transcription start site (TSS) of each target gene of *cis*-eQTL, *cis*-isoQTL, and *cis*-sQTL SNPs. **(F)** Enrichment of *cis*-eQTL, *cis*-isoQTL, and *cis*-sQTLs within functional regions of the genome. **(G)** Loss-of-function mutation intolerance, as measured by pLI score, of eGenes, isoGenes, and sGenes. **(H)** Storey’s pi1 statistic of the proportion of true associations in the discovery group of QTL (y-axis) that are also true associations in the replication group of QTL (x-axis). **(I)** Number of fine-mapping credible sets versus number of conditionally independent eQTLs discovered. The size of the dots is scaled to the number of genes. **(J)** Common recurrent Inversion-QTLs in the developing brain. Inversions are displayed according to their length and the number of overlapping SNPs. Inversions with significantly associated eGenes have filled circles. The size of the circle indicates the population frequency of the inversions.

Compared with adult brain eQTL datasets from PsychENCODE (*13, 14*) and GTEx (*16*), we identified 2,488 unique “fetal-specific” eGenes in this dataset (**Fig. 1B**; **Table S1**). Among conserved eQTL-eGene pairs between the developing and adult human brain cohorts, we observe substantial consistency in effect size as measured by allelic fold change (GTEx: Spearman rho=0.93, p<2.2e-16; PsychENCODE: Spearman rho=0.89, p<2.2e-16; **Fig. 1C**; **Fig. S3**). Notably, these 2,488 fetal-specific eGenes were significantly more intolerant to loss-of-function mutations, as measured by the pLI score (*35*), compared with eGenes shared between fetal and adult timepoints (Wilcoxon p<2e-16; **Fig. S3**), suggesting that these context-specific annotations may be more relevant for interpretation of neuropsychiatric GWAS signals, which are known to be under negative selection. These fetal-specific eGenes were enriched for spliceosomal pathways and cell-type-specific marker genes (*36*) for a subtype of ventricular radial glia (vRG-0; OR 3.3, FDR-corrected P=0.054, Fisher’s exact test; **Fig. S3**; **Table S1**).

Dysregulation of RNA splicing has been strongly implicated in complex disease risk (*29*) as well as alterations in brain development (*24*). To systematically investigate RNA splicing regulation in the developing human brain, we used two complementary approaches. First, guided by existing GENCODE reference transcriptome annotations (*37*), we imputed isoform expression from short-read RNA-seq using Salmon (*38*). Second, we profiled local alternative splicing events via LeafCutter (*39*), an annotation-free method that quantifies intron excision ratios. Next, we identified genetic variants that are significantly associated with these isoform- and splicing-level quantitative traits (**Fig. S4**; **Fig. S5**; Methods), identifying 11,845 and 7,490 genes with a significant *cis* isoQTL or sQTL (termed “isoGenes” and “sGenes”; **Table S1**), respectively. Among the identified xQTL containing-genes, 3,740 are shared, and 2,312/1,891 only have significant genetic regulation detected at isoform/local splicing levels (**Fig. 1D**). As expected, *cis*-xQTL are strongly clustered near their target genes’ transcription start site (TSS) (**Fig. 1E**) and are correspondingly enriched in functional regions of the genome (**Fig. 1F**; **Table S1**). In addition, we observed that isoGenes and sGenes are significantly more intolerant to loss-of-function mutations than eGenes (**Fig. 1G**). To investigate the variant-level overlap among *cis*-e/iso/sQTLs, we calculated Storey’s pi1 statistic (*40*) and found that genetic variants directly associated with RNA splicing (especially isoQTL) capture independent and additional signals from eQTLs (**Fig. 1H**; Methods). Altogether, these results highlight the importance of studying regulation at the level of isoforms and splicing.

We next sought to characterize the extent of allelic heterogeneity in the developing human brain, in which multiple causal variants regulate gene expression at a given locus. Through stepwise conditional QTL mapping (*41*) (**Fig. S3**; **Table S1**; Methods), we identified multiple independent regulatory signals for 3,570 eGenes in the fetal brain, with some exhibiting up to 10 groups of conditionally independent *cis*-eQTL signals. As expected, the primary *cis*-eQTL signal (median - 0.0340 kb) was closer to the corresponding target gene’s TSS than was the secondary *cis*-eQTL signal (median 0.4820 kb; P<2.2e-16, Wilcoxon rank sum). The same was also found to be true for tertiary and higher-rank *cis*-eQTL signals. As an orthogonal approach, we also performed statistical fine-mapping with SuSiE (*42*), which estimates credible sets (CS) of causal variants. The number of SuSiE-estimated CS was highly concordant with the number of conditionally independent QTL signals for each eGene (Spearman correlation 0.66, p<2.2e-16; **Fig. 1I**), indicating that fine-mapping is a complementary approach to conditional QTL mapping for detecting independent regulatory signals. Each CS prioritized a median of 5 SNPs, with 2,423 containing exactly one SNP, which are strong candidates for functional validation (**Data S1**).

Compared to SNPs and indels, the impact of other classes of genetic variation (e.g., structural variants) on downstream gene expression remains underexplored. Nevertheless, complex structural variants such as large recurrent inversions have known associations with brain-relevant traits and can impact gene expression extensively (*25, 43*). To address this, we imputed the genotypes of 17 common (MAF > 0.05) inversions into our uniformly processed fetal brain data and quantified their effects on gene expression across the transcriptome. Longer inversions are more likely to significantly affect downstream gene expression. We found 49 inversion-associated expression quantitative loci (Inv-eQTLs) both in *cis* and in *trans*, for four of the inversions located at 17q21.31, 16p11.2, 7q11.22 and 8p23.1 (**Fig. 1J**; **Fig. S6**; **Table S1**; Methods).

### Cross-Ancestry Gene Regulation and Fine-mapping

Differences in genetic variation (e.g., allele frequency and LD) across ancestries (*44*) has the potential to increase power for statistical fine-mapping (*45, 46*). After filtering and imputing the genotype data (**Fig. S2**; Methods), we inferred genetic ancestries in the fetal brain samples in reference to the 1000 Genomes (*44*). Of the 654 samples, 292 (44.6%) were labeled as “European” (EUR), 164 (25.1%) as “Hispanic/Latino” (AMR), 145 (22.2%) as “African” (AFR), 29 (4.4%) as “Southeast Asian” (SEA), and 24 (3.7%) as “East Asian” (EA) (**Fig. 2A**; **Fig. S1**; **Table S1**). Following the multi-ancestry QTL mapping pipeline, we independently mapped xQTLs in the three largest ancestry groupings: EUR, AMR, and AFR. 986 eGenes were shared between the multi-ancestry dataset and the three ancestry groupings (**Fig. 2B**; **Table S2**). Among the shared eGene-eQTL pairs between ancestries, we observe a highly consistent effect size measured by allelic fold change (*47*) (AMR-EUR Spearman correlation=0.97, p<2.2e-16; AFR-EUR Spearman correlation=0.97, p<2.2e-16; **Fig. 2C**). We then performed statistical fine-mapping separately in the ancestry groupings (*42*), and show that AFR, despite of the smallest sample size, has on average the smallest credible sets (CS) of causal variants (**Fig. 2D; Data S2, S3, S4**). Next, we found that the multi-ancestry dataset substantially reduced the CS size. Taking gene *MTFR1* as an example (**Fig. 2E**), we show that the gain in fine-mapping resolution is not solely due to larger sample size, but the potential of multi-ancestry data to leverage distinct patterns of LD. *MTFR1* has complex LD structure in EUR and AMR, reflected in its large CS (EUR n=49, AMR n=57), whereas in AFR, with simpler LD, the CS is downsized to 9 variants. CS sizes are further reduced in multi-ancestry fine-mapping (n=5), and in a *trans*-ancestry fine-mapping framework that leverages functional annotation (*48, 49*) (n=3). With these results, we highlight the promise of multi-ancestry data in refining statistical fine-mapping.

**Fig. 2.**
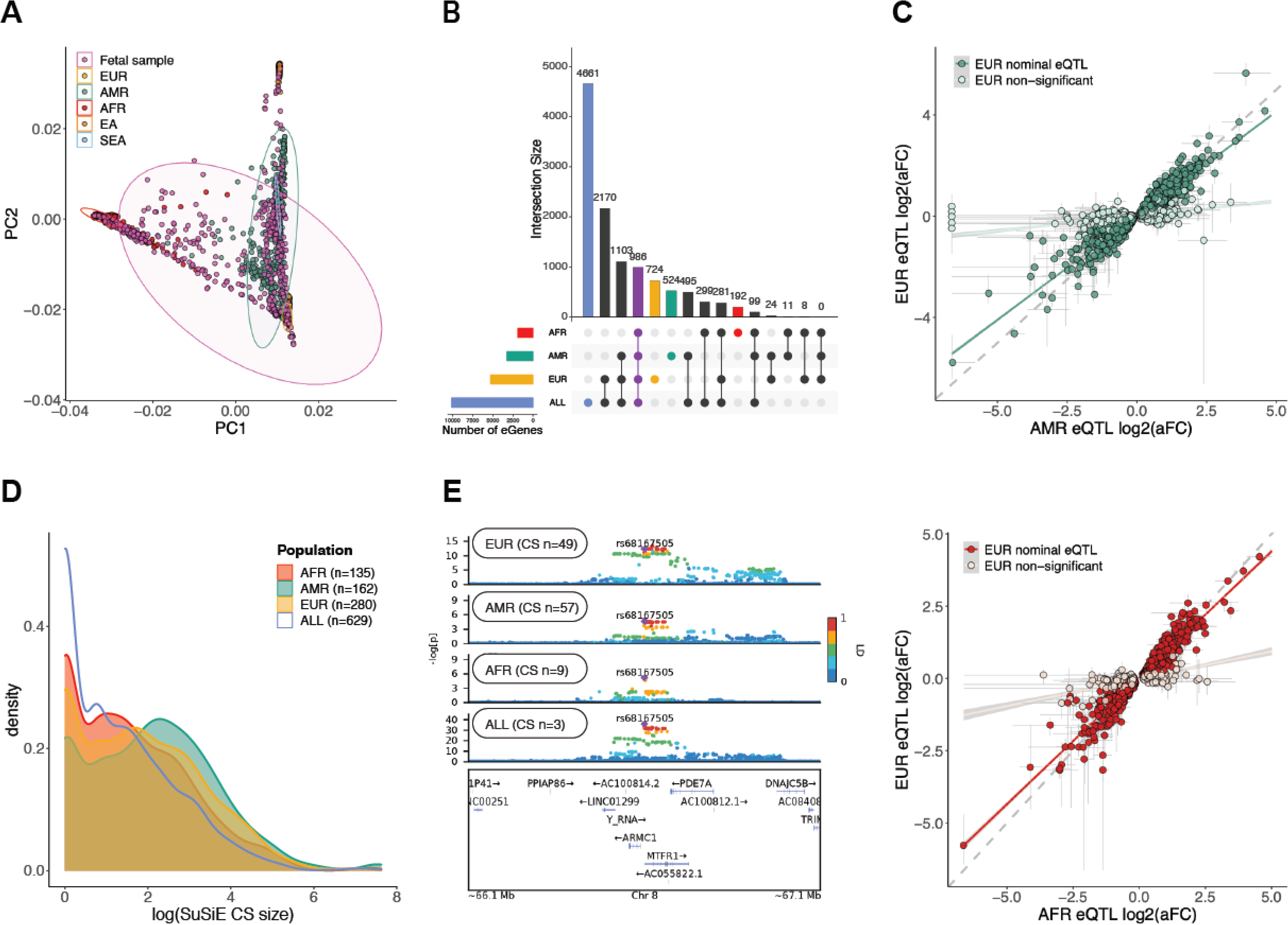
Cross-population gene regulation and fine-mapping. **(A)** Genotype PCA of the fetal brain samples. Population structure was inferred by merging imputed genotypes with 1000 Genomes. **(B)** Comparison of eGenes discovered in the full multi-population dataset (“ALL”, n=629) and in the separate sub-populations, EUR (n=280), AMR (n=162), and AFR (n=135). **(C)** Correlation of eQTL effect sizes between AMR/AFR (top/bottom) and EUR, as measured by allelic fold change. Each dot is an AMR/AFR eGene-primary eQTL pair and colored by its nominal significance in EUR. Grey lines denote the lower and upper bounds of aFC. **(D)** Comparison of fine-mapping credible set sizes between the populations. **(E)** *Cis* associations for the gene *MTFR1* in the cross-population, EUR, AMR, and AFR datasets.

### Trimester-Specific Genetic Regulation

Previous work has implicated the mid-fetal period of human brain development as a critical window convergently impacted by multiple distinct rare genetic risk factors for ASD and SCZ (*21–23*). Although common variant-mediated gene regulation is also known to be dependent on developmental context, this has not been systematically evaluated in the developing human brain. To address this critical gap, we conducted trimester-specific analyses to investigate the specificity of genetic regulation during distinct periods of brain development. We called QTLs after first separating samples into equally powered trimester 1 (4-13 PCW; EUR, n=143) and trimester 2 (14-26 PCW; EUR, n=145) windows, although there were too few samples in trimester 3 (n=4) to conduct a similar analysis. Surprisingly, we identified almost two-fold more trimester 1 eGenes (‘Tri1’; n=4,211) than Tri2 (n=2,220), with 1,261 eGenes shared between the two trimesters (**Fig. 3A-B; Fig. S7; Table S3**). To further explore this unexpected result, we next calculated the *cis*-window SNP-based heritability (*cis*-*h*^2^_SNP_) for gene expression -- a related measure that is more robust to sample size variation -- in Tri1 and Tri2 samples, as well as in adult samples from PsychENCODE (n=1,387). Mirroring eQTL results, we found that *cis* heritability drops significantly from Tri1 to Tri2 (**Fig. 3C**; Wilcoxon p < 2.2e-16; **Table S3; Data S5, S6**). Remarkably, we further observed significantly higher *cis*-*h*^2^_SNP_ in both Tri1 and Tri2 compared with adult brain samples (Tri1 vs adult Wilcoxon p = 7.8e-8; Tri2 vs adult Wilcoxon p < 2.2e-16) (**Fig. 3C**).

**Fig. 3.**
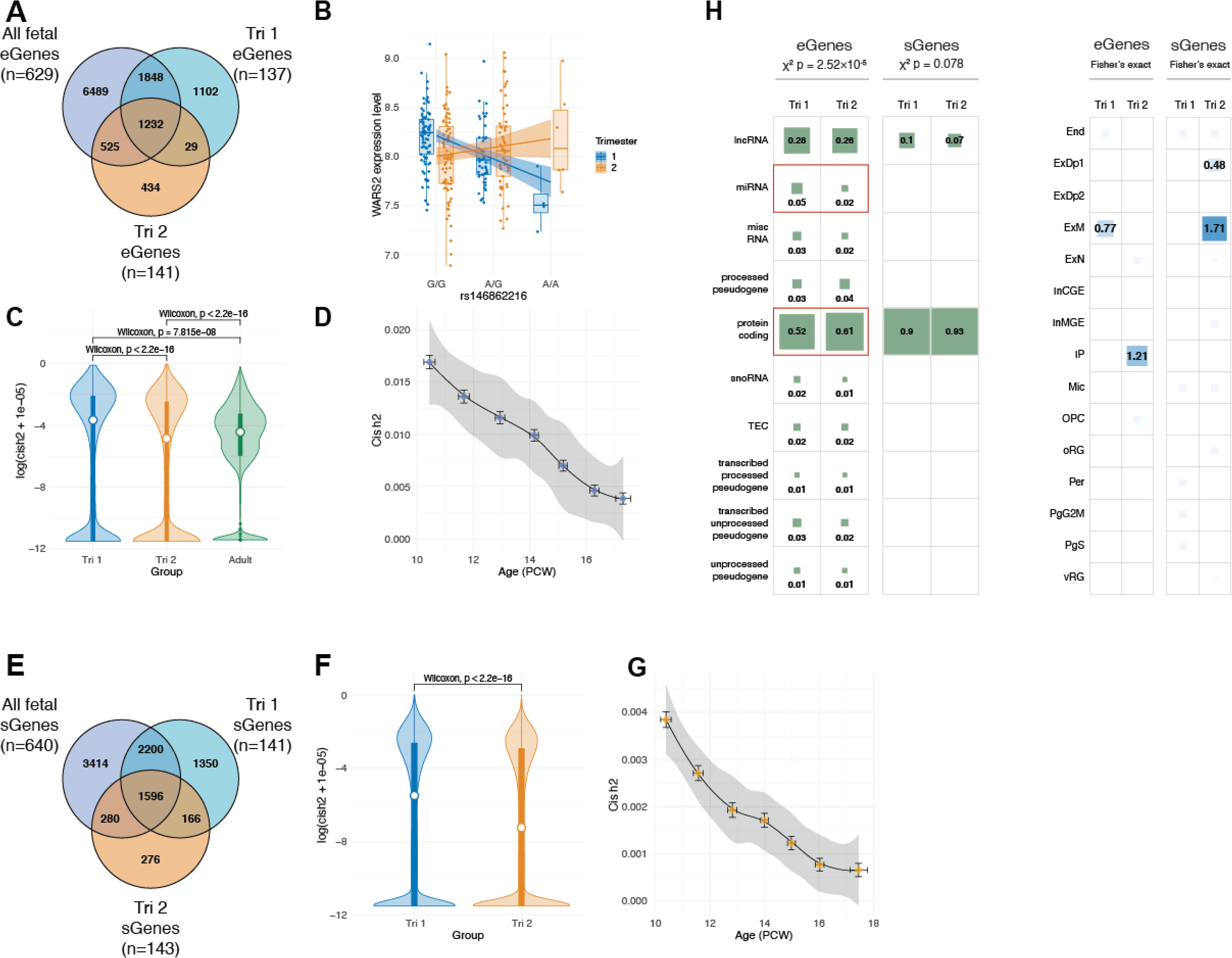
Trimester-specific patterns of gene expression and splicing regulation. **(A)** Comparison of eGenes discovered in Tri1, Tri2, and the full dataset. Notably, we identify many more eGenes in Tri1 than Tri2 despite similar sample sizes. **(B)** Trimester-specific eQTLs for *WARS2*, where rs146862216 (G>A) is an eQTL in Tri1 (beta=-0.89, FDR=3.88e-13) but not in Tri2 (beta=-0.03, P-value=0.71). **(C)** *cis*-heritability of gene expression drops from Tri1 to Tri2 timepoints, and between fetal and adult (PsychENCODE) samples. **(D)** *cis*-heritability of gene expression across 10-18 post-conception weeks (PCW) in fetal brain EUR samples. For D and G, each dot represents a sliding set of temporally ordered samples (n=150), with mean age (+/- SD) on the x-axis, and median *cis*-h^2^_SNP_ (+/- SD) on the y-axis. **(E)** Comparison of sGenes discovered in Tri1, Tri2, and the full dataset. **(F)** *cis* heritability of intron excision ratios in Tri1 vs Tri2. **(G)** *cis* heritability of local splining in fetal brain EUR samples over development, as in (D). **(H)** Comparison of gene and cell type enrichment of Tri1-only and Tri2-only e/sGenes.

We next sought to determine whether a particular developmental inflection point could contextualize this drop in heritability. To address this, we rank ordered EUR samples by PCW, and performed a sliding window analysis of *cis*-*h*^2^_SNP_ across development in equally powered batches (n=150; **Fig. 3D**). We again observed a striking linear drop in *cis*-*h*^2^_SNP_ across the entire range from 10-20 weeks. There were no detectable differences in the number of conditionally independent eQTLs (**Fig. S7**), gene expression levels, or phenotypic variance across Tri1 and Tri2 that could be driving these results (**Fig. S8**). To investigate other potential biological explanations for this drop, we next assessed whether Tri1 or Tri2 eGenes were differentially enriched among specific gene bio-types, cell-types, and/or pathways. Notably, from Tri1 to Tri2, we found that eGenes exhibited a significant shift from non-coding to protein-coding biotypes (chi-square=37.1, df=9, p=2.5e-5). Furthermore, Tri2 eGenes were significantly more enriched for deep layer excitatory neurons, a pattern not observed in Tri1 (**Fig. 3H**). Finally, to determine whether these observations extended beyond gene expression, we conducted similar analyses using sQTLs, as local splicing event quantifications are highly distinct from expression. Here again, we observed a striking drop from Tri1 to Tri2 in the number of genes harboring sQTLs, as well as in their *cis*-*h*^2^_SNP_ from 10-18 weeks (**Fig. 3E-G; Fig. S7; Table S3; Data S7, S8**). This pattern also appeared to be biologically driven by a shift from non-coding to protein-coding biotypes, with proportional changes in sGenes approaching nominal significance (chi-square=3.1, df=1, p=0.08) (**Fig. 3H**). Altogether, these results underscore the temporally dynamic, context-specific nature of gene regulation in the human brain.

### Temporal and Molecular Specificity of Neuropsychiatric GWAS

To prioritize the potential underlying biological context(s) through which genetic risk is conferred, we next sought to integrate our expanded set of functional genomic annotations with neuropsychiatric GWAS results. Using the well-powered SCZ GWAS as an example, we observed substantial enrichment of test statistics when subsetting to fetal brain *cis* e-, iso-, and sQTLs compared with background variants (**Fig. 4A**). Using stratified LD-score regression (S-LDSC (*6, 50*)), we next sought to estimate the degree to which SNP-based heritability was enriched among these xQTLs. We generated continuous annotations for genetic variants based on fine-mapping posterior probabilities (*51*) and found SCZ GWAS signals to be highly enriched among fetal brain regulatory elements -- more so than in the adult brain (**Fig. 4B**; **Table S4**; Methods). When jointly modeling fetal brain e-, iso-, and sQTL, we observed that splicing and isoform QTLs captured greater enrichment than eQTLs (**Fig. S9**; **Table S4**; Methods). These observations were consistent when extending across multiple neuropsychiatric GWAS as well as within trimester-specific annotations (**Fig. S9**; **Table S4**). Overall, these analyses highlight the critical importance of the fetal brain context -- as well as splicing and isoform regulation -- for interpreting potential mechanisms underlying psychiatric GWAS signals.

**Fig. 4.**
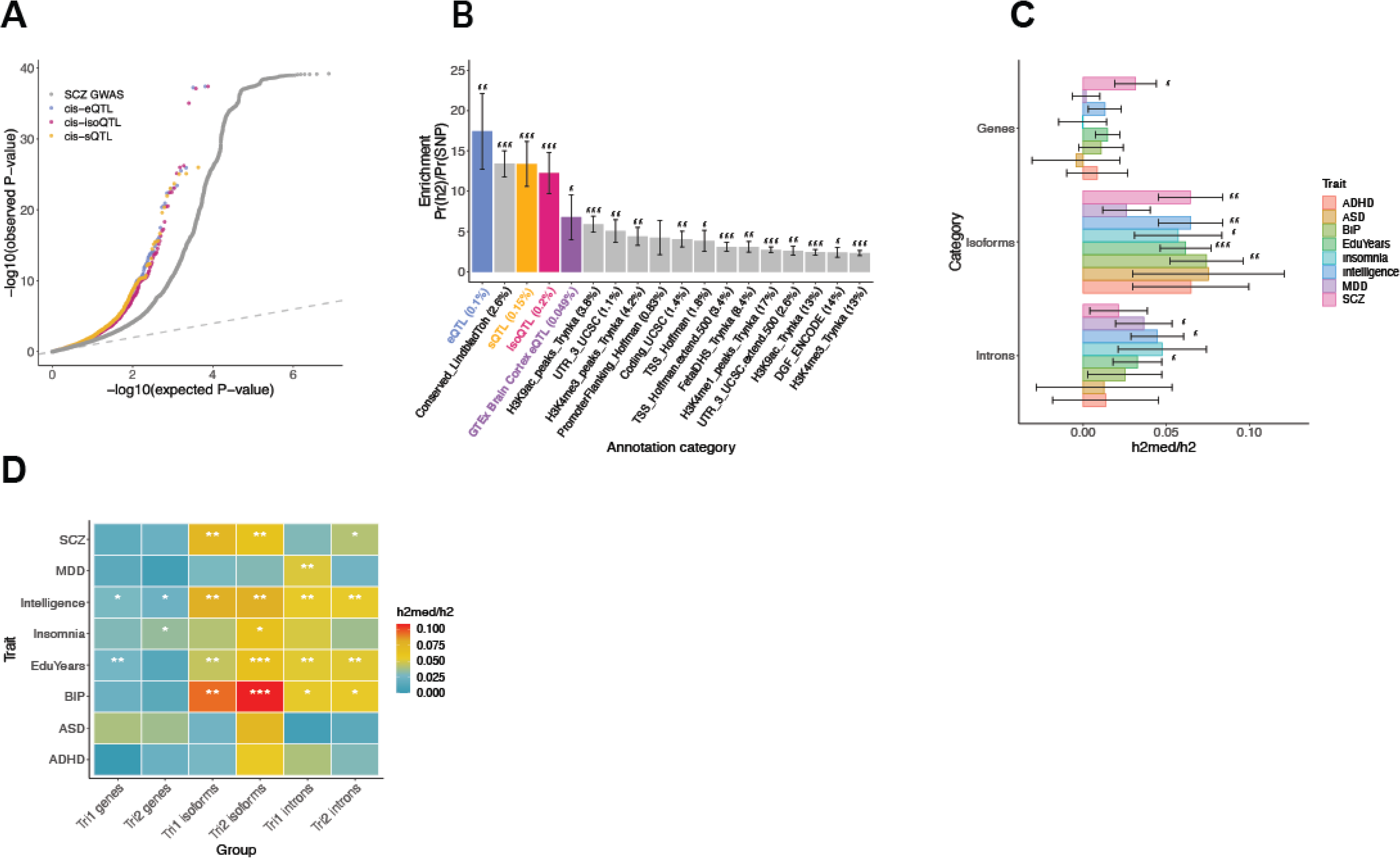
Integrative analysis of fetal xQTLs with neuropsychiatric GWAS. (**A**) Quantile-quantile plot of SCZ GWAS p-values, subsetted by top *cis*-eQTLs, *cis*-isoQTLs, and *cis*-sQTLs in comparison to all background GWAS SNPs. (**B**) s-LDSC enrichment of SCZ GWAS heritability within fetal brain xQTLx, adult brain cortex eQTLs (GTEx v8), compared with background functional annotations. The proportional genomic coverage of SNPs within each annotation are shown in parentheses. For B, C, D: *** FDR<0.001, ** FDR<0.01, * FDR<0.05. (**C**) Estimated proportion (+/- SE) of GWAS *h*^2^_SNP_ mediated by the *cis* genetic component of gene, isoform, and intron (splicing) regulation. Isoform-level QTLs mediate the greatest degree of heritability for multiple neuropsychiatric traits in the developing brain, compared with e/sQTLs. (**D**) Estimated proportion of GWAS *h*^2^_SNP_ mediated by the *cis* genetic component of trimester-stratified gene-, isoform-, and intron (splicing)-regulation.

Although the S-LDSC results clearly demonstrate greater enrichment of SNP-heritability among prenatal (relative to postnatal) gene regulatory variants, and splicing/isoform regulation (relative to total gene expression), these analyses are based on small genomic windows centered around top xQTLs. It can be difficult to compare results across annotations that differ substantially in genomic coverage, and it has been hypothesized that the top (e.g., most heritable) xQTLs may not overlap strongly with complex traits under negative selection (*52, 53*). To address these issues, we next leveraged the mediated expression score regression (MESC) framework (*53*) to estimate the proportion of heritability that is mediated by the *cis*-genetic component of assayed genes, isoforms, and introns (h^2^_med_/h^2^_g_). Notably, while S-LDSC is restricted to subset of significant xQTLs, MESC estimates enrichments genome-wide, including assayed features with low expression heritability (*53*). We chose five GWAS of brain-relevant neuropsychiatric traits SCZ, ASD, bipolar disorder (BIP), attention deficit/hyperactivity disorder (ADHD), and major depression disorder (MDD) (*1, 2, 54–56*). Across these traits, we consistently observed greater heritability mediated by isoform regulation (isoform h^2^_med_/h^2^_g_ 6.45±1.92%) than that of gene expression (3.15±1.25%) or splicing (2.13±1.74%), although the overall extent of mediation remains small (**Fig. 4C**; **Table S4**; Methods). Extending these analyses across trimester-specific annotations, although less well powered, pointed to isoform-regulation in Tri2 as being particularly important, especially for BIP (**Fig. 4D**; **Table S4**). Altogether, our results highlight the importance of fetal isoform expression regulation as a critical mediator of psychiatric GWAS heritability, and the second trimester as the time frame during which genetic risks converge in the developing human brain.

### Colocalization and Isoform-level Transcriptome-wide Association Study (isoTWAS)

To move from broad GWAS enrichments to locus-specific risk genes and molecular mechanisms, we next conducted a systematic colocalization analysis using eCAVIAR (*9*), which performs joint probabilistic fine-mapping and estimates the colocalization posterior probability (CLPP) that a given variant is “causal” in both GWAS and xQTL datasets. Across the combined 485 psychiatric GWAS loci, we are able to prioritize 292 (60.2%) with CLPP above the established cutoff of 0.01 (**Fig. 5A**, **B**; **Fig. S10**; **Table S5**; **Methods**). For context, restricting our results to eQTLs in SCZ and BIP, we identify 80 loci with prioritized eQTLs, while a recent, much larger adult brain eQTL study with an effective sample size of 3,154 identified 20 significant colocalizations (CLPP>0.01) for the two disorders (*15*).

**Fig. 5.**
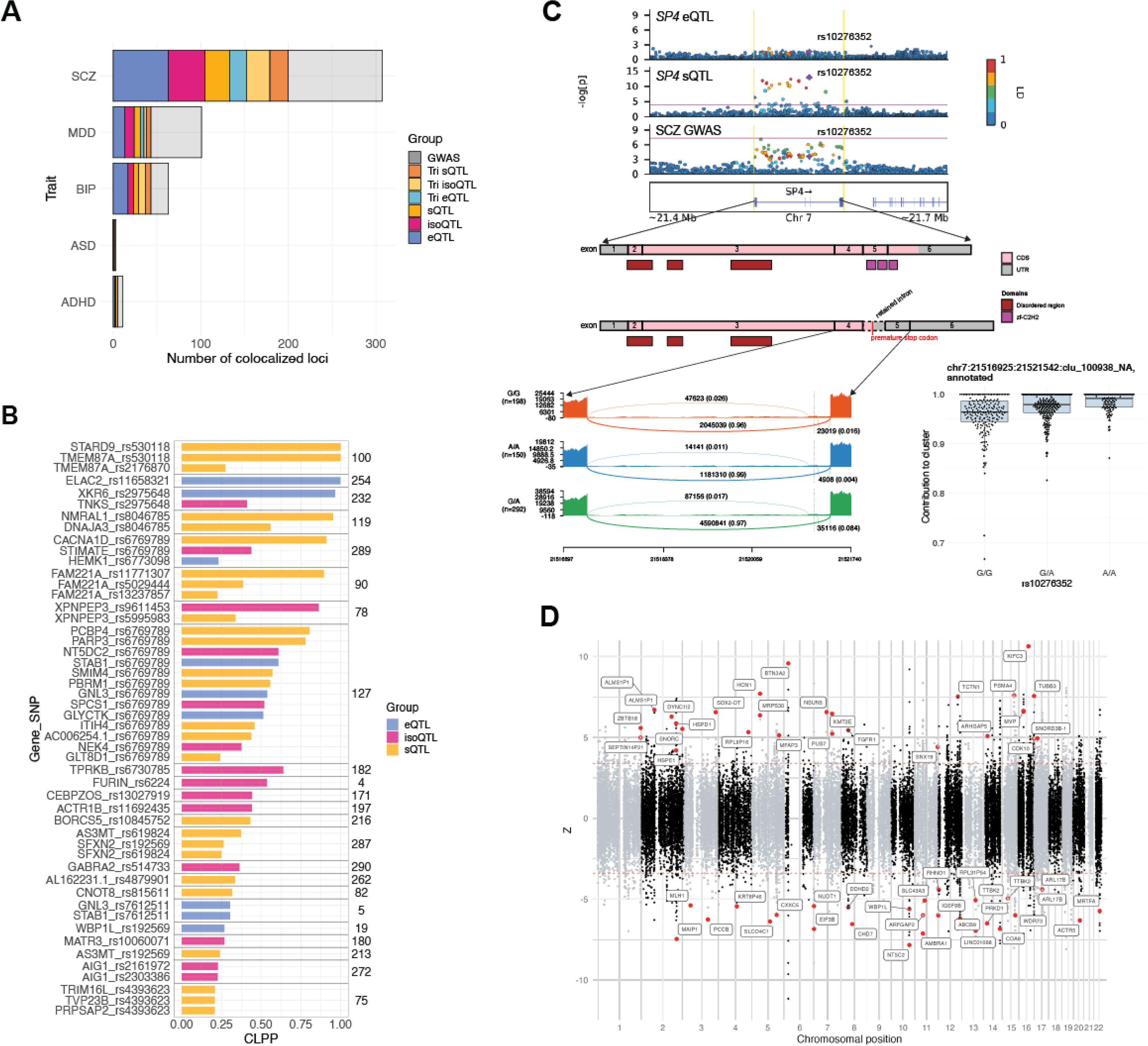
Neuropsychiatric risk gene prioritization through colocalization and isoTWAS. (**A**) Total number of GWAS loci exhibiting significant colocalization (CLPP>0.01) with specific fetal brain xQTL annotations. From left to right, bars represent the cumulative number of GWAS loci colocalized with each additional annotation. Shown in grey are GWAS loci that are not colocalized with any annotation. (**B**) Colocalization between SCZ GWAS and fetal brain xQTLs, ranked by CLPP and grouped by GWAS locus, as indicated by the number to the right. (**C**) Top: locus plots of SCZ GWAS with *SP4* e- and sQTLs. A significant colocalization is observed for SCZ GWAS with a cryptic splicing event in *SP4*. Notably, *SP4* does not have a detectable eQTL in the fetal brain. Middle: Gene structure of *SP4* with and without cryptic exon inclusion, likely resulting in nonsense mediated decay. Bottom left: sashimi plot shows the density of exon and junction read mapping for intron cluster chr7:21516925-21521542, stratified by the colocalized sQTL. Bottom right: sQTL rs10276352 (G>A) increases the contribution to cluster of annotated intron chr:21516925-21521542. The SCZ risk allele increases cryptic exon inclusion. (**D**) Fetal brain isoTWAS associations with SCZ GWAS. Each dot represents an isoform and genes with significant, prioritized isoforms are colored in red.

Consistent with heritability enrichment patterns, we observed many more GWAS loci harboring colocalized iso-QTLs and sQTLs, compared to standard eQTLs (**Fig. 5A**,**B**; **Fig. S10**; **Table S5**). As a validation, we recover the well-established neuropsychiatric risk gene *FURIN*, with an isoQTL colocalization observed across both SCZ and BIP (**Fig. 5B**; **Fig. S10**) (*10*). The colocalized variant rs6224 (located at intron 13) is in moderately strong LD (R2=0.7) with rs4702 (located at the 3’-UTR of *FURIN*) that has been CRISPR-validated as an eQTL (*57, 58*). Further, we uncover a common-variant mechanism underlying the SCZ GWAS association at *SP4*, a transcription factor with a high-confidence rare variant implicated SCZ risk gene (*1, 59*). While *SP4* was known to be the target gene in this GWAS locus, variant-to-gene mapping was limited by the lack of an observable *SP4* eQTL signal in adult or fetal brain. Here, we identify a novel cryptic splicing event in *SP4* colocalizing with both SCZ and BIP GWAS (**Fig. 5C**; **Fig. S10**). The risk variant rs10276352 (G>A) was identified as an sQTL associated with increased inclusion of a 181 bp cryptic, unannotated exon (chr7:21521120-21521300). The inclusion of this cryptic exon is predicted to introduce a frameshift and premature stop codon between canonical exons 4 and 5, likely resulting in nonsense-mediated decay. The resulting truncated protein, if any, would be missing the zinc finger domain that is critical for *SP4*’s DNA-binding activity (**Fig. 5C**).

Other novel disease mechanisms prioritized by these analyses include: downregulation of the GABA_A_-receptor subunit *GABRA2* in SCZ (eQTL: rs514733, CLPP=0.32; **Fig. 5B**; **Table S5**), providing genetic evidence for GABAergic dysfunction hypothesized to contribute to disease pathophysiology (*60*); splicing dysregulation of *ADCY2* in BIP (top colocalization sQTL: rs11750832, CLPP=0.97; **Fig. S10**; **Table S5**) which encodes adenylate cyclase, a cell membrane-bound enzyme regulating cAMP signaling and a target of the mood stabilizer lithium (*61*); splicing dysregulation in BIP of *SCN2A* (sQTL: rs17183814, CLPP=0.70; **Fig. S10**; **Table S5**), a voltage gated sodium channel subunit with rare variant associations in ASD and epilepsy (*62, 63*); and in MDD, the dysregulated splicing of *CBLL1* (CLPP=0.43; **Fig. S10**; **Table S5**), a highly constrained E3 ubiquitin-protein ligase and regulator of N6-methyladenosine RNA modification involved in neural-immune activation (*64*).

We next conducted an isoform-centric transcriptome-wide association study (isoTWAS) (*65*), to identify genes and their isoforms whose *cis* regulated expression is associated with SCZ risk. isoTWAS identified 536 isoforms across 271 unique genes with significant isoTWAS associations (Bonferroni-corrected P < 0.05 across genes, FWER-corrected P < 0.05 across transcripts of the same gene, permutation P < 0.05; Methods). To leverage local LD and to account for SNP weight correlations, we performed fine-mapping on isoTWAS associations that passed permutation testing (**Fig. 5D**; **Table S5**). This analysis resulted in 129 putatively causal isoforms from 107 unique genes that fall in 90% credible sets, with 22 of these genes with pLI score > 0.8. Of these, 69 isoforms from 57 unique genes are within 500 kb of a GWAS-significant risk SNP. In addition, among LD-independent blocks that have SCZ GWAS significant SNPs, we identified 41 independent GWAS SNPs within 500 kb of an isoTWAS-prioritized isoform. Comparing isoforms prioritized through isoTWAS and/or isoQTL colocalization revealed 14 isoforms that were prioritized with both methods, including *SLC9C2, KMT2E,* and *ABCB9.* Of note, the relatively low overlap could be due to the low power of probabilistic colocalization methods (*66*). With its increased power, isoTWAS also captured additional notable associations, including genetically mediated upregulation of *HCN1-201*, the dominant isoform of the hyperpolarization-activated cation channel *HCN1*. An additional advantage of isoTWAS over colocalization analyses is the ability to map isoform-level associations outside of GWAS-significant loci, additionally identifying putatively causal 60 isoforms across 50 unique genes that were found outside a GWAS-significant locus.

When examining results from gene prioritization analyses, we noticed several instances in which a single variant was associated with multiple distinct QTLs and significantly overlapped with disease GWAS signal. For example, rs6769789 was identified as an xQTL for multiple genes on chromosome 3 that colocalized with SCZ GWAS (**Fig. 5B**). Due to pleiotropy, it can be particularly difficult to distinguish whether one (of many potential) molecular effects is the true mediator of the SNP-trait effect. In attempt to address this, we employed MRLocus (*67*) which leverages a Mendelian randomization framework for loci with allelic heterogeneity, using conditionally-independent QTLs for these mediators to further support (or refute) observed gene-to-trait effects. Among 14 TWAS-identified genes with >3 conditionally independent eQTLs, the MRLocus framework provided additional support for 7 genes with FDR<0.2 (**Table S5**; Methods). In the pleiotropic rs6769789 locus described above, for example, there was an association between eQTL and GWAS effect sizes among conditionally independent eQTLs for *NT5DC2* (FDR=0.125), providing additional evidence in support of this gene-trait association (**Fig. S11**).

### Network-level Contextualization of Developmental Gene Regulation

To identify developmentally relevant transcriptional networks, interrogate them for common and rare genetic risk, and connect that risk with key biological and cellular processes, we performed robust weighted gene co-expression network analysis (rWGCNA) on our dataset, an unsupervised method that clusters genes into modules based on shared patterns of expression (*22, 68–70*). We constructed networks using gene and isoform-level quantifications across all samples (n=642), as well as within samples filtered by trimester (Tri1, Tri2) or chromosomal sex (XX, XY). In total, we identified 124 gene and isoform co-expression modules demonstrating enrichment for all major developmental cell types (*36*) and recapitulating early biochemical processes and developmental pathways (**Fig. 6A**; Methods). These networks expand on previous work (*12, 18, 19, 21, 22, 24*) by incorporating an order of magnitude greater number of samples, generating isoform-level modules for over 120,000 transcripts, and contextualizing development-wide findings within trimester- and sex-specific contexts.

**Fig. 6.**
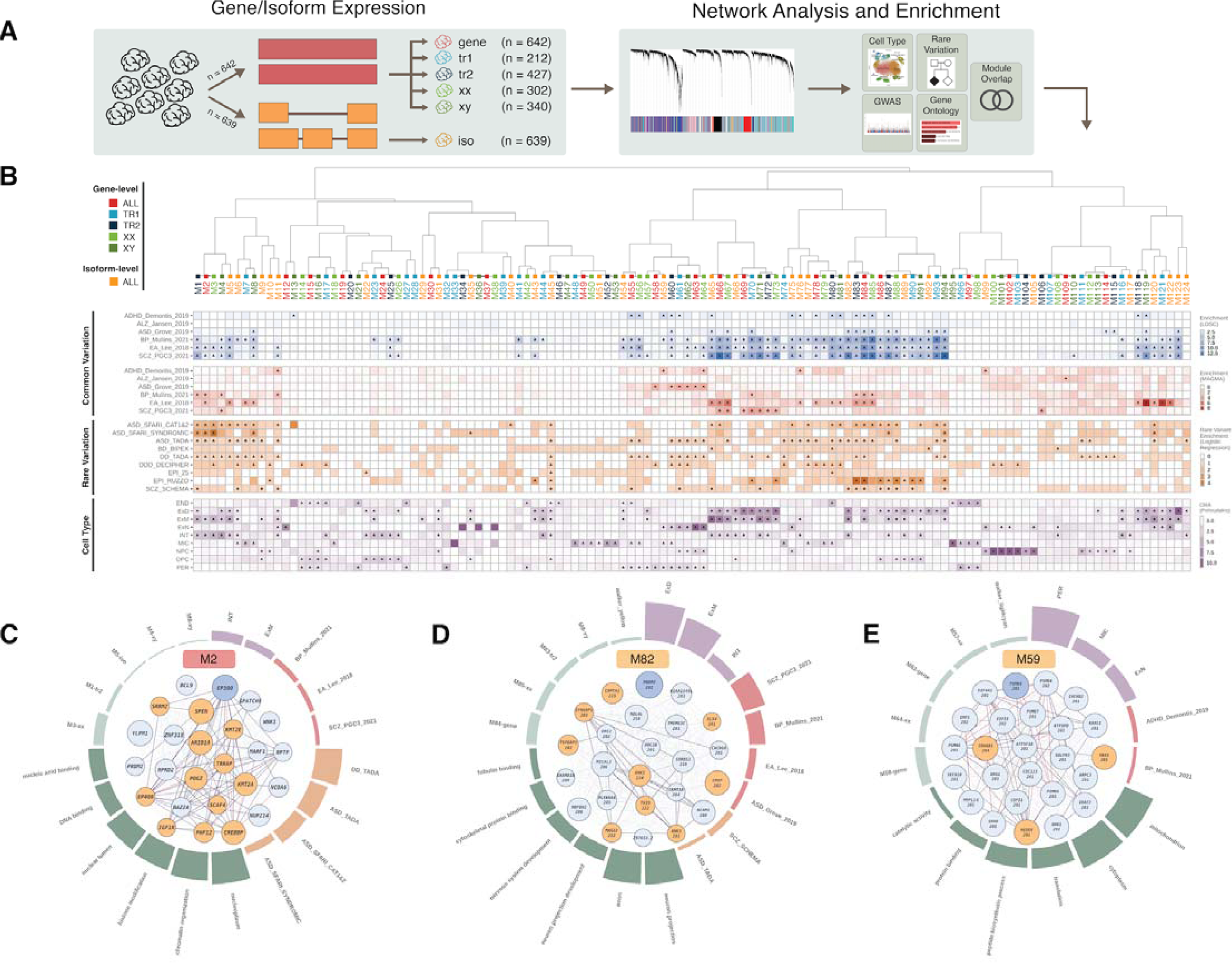
Systems-level integration of risk variation with developmental gene and isoform co-expression. (**A**) Workflow for construction of gene and isoform-level co-expression networks, followed by cell-type, pathway, and disease gene enrichment analyses. Separate gene co-expression networks were built to capture trimester and sex-specific effects. (**B**) Top: hierarchical clustering of modules from gene, isoform, trimester and sex-stratified networks through bi-weight mid-correlation of eigengenes. Middle: heatmaps depict module-level enrichment for neuropsychiatric GWAS signal (-log_10_P_enrich_ from s-LDSC and MAGMA) and odds ratios (OR) for rare variation and cell type enrichment (truncated at 10). Triangles indicate FDR-corrected P<0.1 significance. (**C**) Annotations for M2, a development-wide disease-associated chromatin regulation module. Center: top module (‘hub’) genes with circle size reflecting module membership (kME) and orange shading indicating genes with associated high-confidence neuropsychiatric disorder associated rare variants. Thin edges represent topologic overlap, solid edges indicate protein-protein interactions from the STRING database. Surrounding: circular bar plot highlighting module enrichment for cell types (purple), common (red) and rare (red-orange) variation, gene ontology terms (dark green), and module overlap (light green). (**D**) Annotations for M82, a SCZ/BIP-associated deep layer neuron-projection isoform module. (**E**) Annotations for M59, an associated mitochondrial/proteasome isoform module.

Overlaying enrichment for common and rare variation within the hierarchical structure defined by co-expression pinpointed groups of modules that harbor most of neurodevelopmental disease risk (**Fig. 6B**; **Table S6**; Methods). Broadly, we find that isoform modules are more likely to capture cell-type marker enrichment (two-sided Fisher’s exact test on FDR<0.1 hits, p=8e-5, OR=2.37), demonstrating the importance of incorporating splicing and isoform expression in a network context. Neuropsychiatric GWAS signals were found to localize within modules enriched for deep layer excitatory, maturing excitatory, and interneuron cell populations, while exhibiting depletion for neural progenitor modules. Rare variation was more likely to be enriched in deep layer excitatory, maturing excitatory, and oligodendrocyte precursor modules, suggesting that the majority of disease-associated variation perturbs the maturation of neuronal cell types (**Fig. S12**).

At the gene level, a group of modules (M1, M2, M3) enriched for chromatin remodeling and histone modification pathways, with hub genes including *EP300*, *EP400*, *ARID1A*, *KMT2E* and *POGZ*, converged with excitatory and inhibitory neuron marker genes and showed strong enrichment for rare variation associated with ASD and developmental delay (DD) (**Fig. 6C**). These modules highly overlapped with the DD- and ASD-risk yellow module described in Walker et al. (*24*), building upon the previously described excitatory neuron enrichment for these disorders. Remarkably, our analysis partitions the hub genes of the yellow module into another group of excitatory-enriched synaptic projection modules (M83, M84, M85; **Fig. S12**), which exhibits strong enrichment for cross-disorder common variation and rare variation risk for ASD, epilepsy, and SCZ. Among the strongest genetic risk for SCZ was observed in two groups of interrelated synaptic modules (group 1: M93, M94; group 2: M86, M87, M88; **Fig. S12**), which strongly enriched for synaptic gene ontologies and excitatory neuron marker genes. A closely related isoform module, M82, uniquely captured this SCZ signal and harbored hub transcripts with known risk, like the canonical transcripts of *TRIO* and *SYNGAP1*, as well as multiple transcripts of *ANK3* (**Fig. 6D**). Using module preservation analysis, we identified M17 as a first trimester-specific module that shows unique enrichment for neurodevelopmental disorders (NDD) risk genes and ontologies for neurogenesis and DNA binding, with hub genes such as *GREB1L*, *LRP2*, and *CASZ1* (**Fig. S12**).

At the isoform level, M59 stood out for its consistent and unique association with ADHD common variant risk and GO term enrichment for mitochondrial and bioenergetic pathways. Hub genes for this module included multiple proteasome subunit genes (PSMD family), and cell type analysis revealed association with excitatory, microglial, and pericyte populations (**Fig. 6E**). Furthermore, a group of neuronally-enriched modules (M65, M66, M67; **Fig. S12**) displayed robust BIP, education attainment (EA), and SCZ enrichment while harboring high risk hub genes like *MEF2C* and *SATB2* – similar to the prenatally-enriched M37 module described in Li et al (*18*). Of note, distinct transcripts of known ASD risk gene *SOX5* acted as hub genes for two isoform modules, M120 and M122, but M120 alone showed a strong and specific signal for ASD rare variation (**Fig. S12**).

### Cell-type Specificity through Module-interaction QTLs

Finally, as gene regulation is often known to occur within cell-type-specific contexts, we leveraged two orthogonal approaches to integrate cellular specificity into our gene regulatory results in the developing human brain. First, we applied the network-based framework implemented in CellWalker to integrate fetal single-cell chromatin accessibility with eQTL results (*71*). CellWalker mapped 21.7% of eQTL-containing genes to a specific cellular context in the developing human brain, corresponding to 3,739 of the bulk-derived eQTLs (**Fig. 7A**; **Table S7**; Methods). Second, we leveraged the fact that co-expression modules are known to capture cell-type-specific processes (**Fig. 6A**) to identify module-interacting eQTL (ieQTLs). To identify such ieQTLs, in which SNP-gene associations are modulated by the levels of a given module, we tested the interaction effect between genotype and module eigengenes on gene and isoform expression (**Fig. 7B**; Methods). Across all gene and isoform modules, 8,008 ieQTLs were identified after the permutation test (BH-corrected FDR<0.05; **Methods**), in which 3,960 ieQTLs were specific to a single module (**Fig. 7C**; **Data S9**). To validate the cell-type specificity of module ieQTLs, we examined the sharing between module ieQTLs with eQTLs identified separately within cultured neurons and progenitors (*72*). Using the Storey’s pi1 statistic (*40*), we observed substantial concordance between module interacting SNP-gene pairs that are true associations in external neuron/progenitor eQTL datasets (**Fig. 7C**). 40 modules have a pi1 difference greater than 0.2 between neuron and progenitor eQTLs, suggesting cell-type-specific genetic regulation (**Fig. 7C**). Finally, integrating disease GWAS, we identified significant colocalizations (CLPP>0.01) between ieQTLs and 22 SCZ GWAS loci, 13 of which are not found with bulk *cis*-eQTLs (**Table S5**). For example, we identified a SCZ colocalization with an ieQTL of *BRINP2* and the Tri1 gene module M93 (rs17659437, CLPP = 0.01653) (**Fig. 7D**). M93 is enriched for deep layer excitatory neuron markers (ExD), cross-disorder psychiatric GWAS signal, and synapse-related pathways (**Fig. 7E**), with eigengene expression increasing across development (**Fig. 7F**). Concordantly, this module shows greater pi1 concordance with neuronal compared to progenitor eQTLs (**Fig. 7C**). *BRINP2* is involved in BMP and retinoic acid signaling pathways, both of which play critical roles in brain development and neuropsychiatric disease. In sum, these analyses demonstrate how publicly available data can be used to annotate bulk QTLs to specific cell types, adding further context for detecting gene regulatory variation relevant to neuropsychiatric disease mechanisms in the developing human brain.

**Fig. 7.**
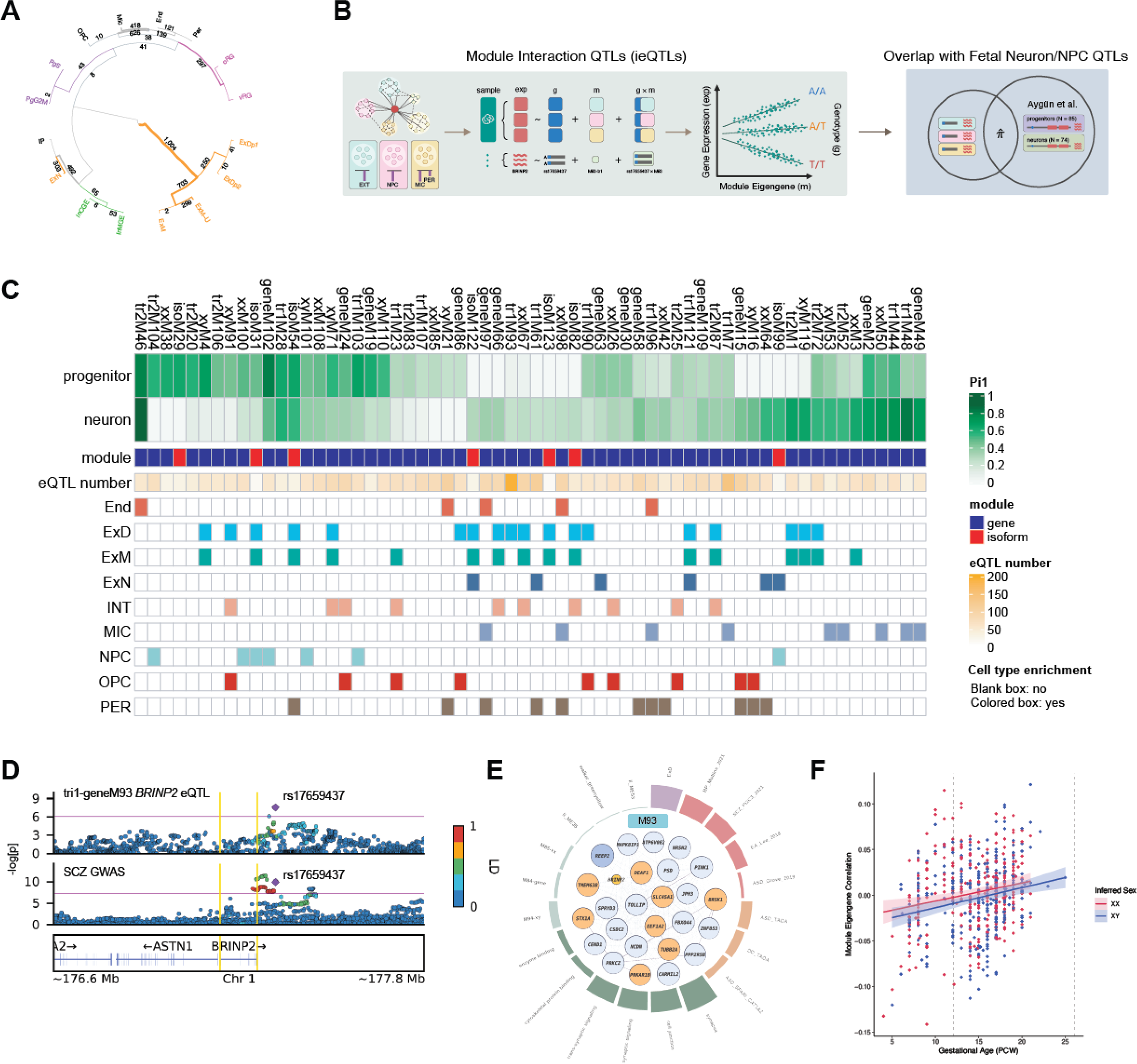
Module interacting eQTLs and context-specific GWAS colocalization. (**A**) Hierarchical clustering of *cis*-eQTL enrichments among specific fetal brain cell-types as mapped by CellWalker. Outermost numbers denote results from single cell type label analysis. Inner numbers denote results from a broader, multi-level label analysis. See Methods for details. (**B**) A schematic of module interaction ieQTL mapping and validation in cultured neurons and progenitors. (**C**) Results from module ieQTL mapping. From top to bottom, pi1 statistics depicting ieQTL overlap with eQTLs from cultured neurons or neural progenitor cells (NPCs), molecular feature (gene/isoform), number of ieQTLs, and cell type enrichment. 62 modules with pi1>0.2 in either neurons/NPCs are shown. (**D**) Colocalization between SCZ GWAS and *BRINP2* ieQTL rs17659437 in M93 (CLPP=0.02). (**E**) Annotation for M93, a SCZ/BIP enriched deep-layer neuronal/synaptic module. (**F**) Trajectory of M93 eigengene expression across brain development, colored by biological sex.

## Discussion

Here, we present an unprecedented view of the landscape of gene, isoform, and splicing regulation in the developing human brain, across more than 650 distinct donors. We further provide xQTL maps specific to the first and second trimesters of human brain development, as well as within 3 genetic ancestries, by leveraging the resulting allelic diversity through *trans*-ancestry fine-mapping to narrow in on underlying causal genetic variants. With this work, we are able to prioritize 2-fold more risk genes and molecular mechanisms underlying neurodevelopmental and psychiatric disorder GWAS loci compared with much larger adult brain reference panels, highlighting the importance of developmental context when interpreting genetic risk variation. Finally, we build gene and isoform-level co-expression networks to place this polygenic risk within an unsupervised, systems-level developmental and cell-type-specific contextualization.

Our results have several notable implications for future investigations of the functional genomic underpinnings of brain-relevant complex traits and disorders. First, several findings highlight the substantial impact of developmental stage on gene regulation, including a striking drop from 10-18 PCW in *cis*-heritability of gene expression and splicing which was not attributable to differences in gene expression magnitude or variance over time. As eQTL discovery and *cis* heritability are interrelated, this likely explains why we are able to identify eGenes on par with larger adult brain studies. Furthermore, while disease genes tend to be under negative selection, which is inversely related to gene expression heritability (*52, 53*), our data suggest that this relationship is likely moderated by developmental stage. Indeed, the fetal brain -- and Tri2, in particular -- harbors greater enrichment for neuropsychiatric GWAS signal than at postnatal timepoints, consistent with results from rare variant enrichment analyses (*21–23*).

Second, we observe striking differences in the ability of distinct molecular features to capture neuropsychiatric risk mechanisms. Specifically, isoform-level regulation mediated substantially more heritability than eQTLs, across several distinct disease GWAS. Further, iso- and sQTL colocalizations explained many more GWAS loci than standard eQTLs from the fetal or adult brain. Given that isoform quantification requires accurate imputation from short-read RNA-seq, guided by genomic reference annotations which are notably incomplete with respect to the human brain (*73*), we expect these differences to only become further accentuated as our understanding of the landscape of alternative splicing and isoform complexity grows, through the continued adoption of emerging long-read RNA-sequencing technologies.

Third, with the ever-increasing number of available functional genomic reference panels, including from this study, it is becoming increasingly clear that there will soon be multiple potential, prioritized (e.g., colocalized) functional genomic mechanisms at a given GWAS locus. This can occur when a GWAS SNP tags a haplotype containing an underlying structural variant (e.g., inversion), with multiple pleiotropic effects on *cis* gene regulation. Mendelian randomization-based approaches can help address this challenge, but will require larger reference panels capable of capturing the full extent of allelic heterogeneity across the transcriptome. Further, while single-cell QTL studies have been heralded of late, they will only exacerbate this issue while lacking the ability to profile splicing and isoform-level regulation which we show to be most high-yield for mechanistic inference.

Finally, given our finding that isoform-level co-expression networks in particular are able to recapitulate cell-type-specific biological processes and genetic regulation, future work should continue to build larger bulk-tissue reference panels of the developing human brain guided by more complete, matching isoform-level annotations from long-read sequencing. Greater visibility is needed, especially in the third trimester of brain development, which is critically underrepresented in our sample.

## Data Availability

Supplemental data can be found on Synapse at https://www.synapse.org/#!Synapse:syn50897018.3/datasets/. Analysis code can be found on GitHub at https://github.com/gandallab/Fetal_metaQTL

## Acknowledgements

We are grateful to the brain tissue donors and their families, without whom this work would not have been possible. This work was supported by the Simons Foundation (SFARI Bridge to Independence Award to M.J.G.), NIMH (R01-MH121521, R01-MH123922 to M.J.G.). Data were generated as part of the PsychENCODE Consortium, supported by: U01DA048279, U01MH103339, U01MH103340, U01MH103346, U01MH103365, U01MH103392, U01MH116438, U01MH116441, U01MH116442, U01MH116488, U01MH116489, U01MH116492, U01MH122590, U01MH122591, U01MH122592, U01MH122849, U01MH122678, U01MH122681, U01MH116487, U01MH122509, R01MH094714, R01MH105472, R01MH105898, R01MH109677, R01MH109715, R01MH110905, R01MH110920, R01MH110921, R01MH110926, R01MH110927, R01MH110928, R01MH111721, R01MH117291, R01MH117292, R01MH117293, R21MH102791, R21MH103877, R21MH105853, R21MH105881, R21MH109956, R56MH114899, R56MH114901, R56MH114911, R01MH125516, R01MH126459, R01MH129301, R01MH126393, R01MH121521, R01MH116529, R01MH129817, R01MH117406, and P50MH106934 awarded to: Alexej Abyzov, Nadav Ahituv, Schahram Akbarian, Kristin Brennand, Andrew Chess, Gregory Cooper, Gregory Crawford, Stella Dracheva, Peggy Farnham, Michael Gandal, Mark Gerstein, Daniel Geschwind, Fernando Goes, Joachim F. Hallmayer, Vahram Haroutunian, Thomas M. Hyde, Andrew Jaffe, Peng Jin, Manolis Kellis, Joel Kleinman, James A. Knowles, Arnold Kriegstein, Chunyu Liu, Christopher E. Mason, Keri Martinowich, Eran Mukamel, Richard Myers, Charles Nemeroff, Mette Peters, Dalila Pinto, Katherine Pollard, Kerry Ressler, Panos Roussos, Stephan Sanders, Nenad Sestan, Pamela Sklar, Michael P. Snyder, Matthew State, Jason Stein, Patrick Sullivan, Alexander E. Urban, Flora Vaccarino, Stephen Warren, Daniel Weinberger, Sherman Weissman, Zhiping Weng, Kevin White, A. Jeremy Willsey, Hyejung Won, and Peter Zandi. Author Contributions: This study was conceived and designed by MJG and CW with input from CL and KSP. Data was provided by DHG, NS, NJB, DRW, AEJ, JEK, TMH, RLW. Analyses were performed by CW, MM, RD, PZ, PFP, DDV, AB, NM, CJ, MK, ET, CH, NA, MIL, supervised by MJG, with contributions from CC, DC, HP, MG, NPD, ZW, KR, AG, BP, MAP, JLS, MIL, KSP, CL. CW and MJG wrote the manuscript, with major contributions from MM, AB and critical input from all authors.

## Competing Interests

M.J.G. and D.H.G. receive grant funding from Mitsubishi Tanabe Pharma America.

## Data Availability

Supplemental data can be found on Synapse at https://www.synapse.org/#!Synapse:syn50897018.3/datasets/

## Code Availability

Analysis code can be found on GitHub at https://github.com/gandallab/Fetal_metaQTL

