## Supplementary Information for "Cross-ancestry, cell-type-informed atlas of gene, isoform, and splicing regulation in the developing human brain"

**The PDF file includes:**

Materials and Methods

Figs. S1 to S12

Captions for Tables S1 to S7

Captions for Data S1 to S9

**Other Supplementary Materials for this manuscript include the following:**

Tables S1 to S7

Data S1 to S9

### Materials and Methods

#### 1: Study descriptions

Here, we integrate the largest-to-date dataset of the developing human brain by combining five individual studies. The Walker (*24*) data was generated at UCLA with 219 brain cortex samples spanning 14-21 post-conception weeks (PCW). For these data, he RNA-seq library was prepared using Ribo-Zero with 60M reads per sample, and genotype data were sequenced using Illumina HumanOmni2.5 array. The O’Brien (*25*) data has 120 bulk brain samples spanning 12-19 PCW, with RNA-seq data having been generated using Ribo-Zero with 100M reads per sample, and genotype sequencing yielded approximately 710,000 SNPs using the Illumina Infinium OmniExpress-24 BeadChip array. The Werling (*12*) data has 116 DLPFC samples spanning 6-37 PCW, with RNA-seq having been generated using Ribo-Zero with 37M reads per sample. There were genotype sequenced using 30X WGS. The LIBD (*26*) data has 54 DLPFC samples spanning 14-20 PCW, with RNA-seq having been generated using Ribo-Zero with 150M reads, and genotype sequencing was performed using Illumina 1M DuoV2. The HDBR (*27*) data has 173 bulk brain samples spanning 4-20 PCW, with RNA-seq data having been prepared using PolyA with 50M reads. For this dataset, genotype sequencing was performed using Illumina HumanOmni5-4v1-B. 10 of the HDBR samples were duplicated in the O’Brien dataset, and these 10 samples were thus removed. Altogether, there are 672 unique samples. We further filtered for matching DNA-RNA and high-quality data and the final sample size is 654 (Walker n=211, O’Brien n=120, Werling n=116, LIBD n=44, HDBR n=163).

#### 2: Genotype processing

Individual study genotype data were uniformly processed. Variants were filtered by MAF, HWE, and missingness (plink --maf 0.01 --geno 0.05 --mind 0.10 --hwe 1e-6(*74*)). Conform-gt (<https://faculty.washington.edu/browning/conform-gt.html>) was applied to genotype data to fix strand flips and to make it consistent with 1000 Genomes Project hg19 reference(*44*). QC-ed genotype data were imputed on the Michigan Imputation Server(*75*) (Minimac 4, phasing: Eagle v2.4) with TOPMed Freeze5(*34*) as the reference, and filtered for high imputation quality (R2 > 0.3). After imputation, individual studies were merged by intersecting the variants. Duplicated position variants (i.e. multi-allelic) were removed. TOPMed variant IDs were mapped to dbSNP151 (UCSC snp151.txt.gz) rsID (~10% variants were not mapped to rsID, and kept as hg38 CHR:POS:REF:ALT as ID). Variants were filtered (plink --maf 0.01 --geno 0.05 --hwe 1e-6) and mapped to hg19 using Crossmap (v0.5.2)(*76*). CheckVCF (<https://github.com/zhanxw/checkVCF>) was used for a sanity check. LD-pruned (plink --indep-pairwise 50 5 0.2) data were merged with 1000 Genomes. PCA and k-nearest-neighbors (k=50) were run to infer data ancestry. Variant QC and PCA were then conducted separately within EUR, AMR, and AFR populations. 8,420,206 variants (including 560,448 indels) were included in the final mixed ancestry data analysis (EUR 6,381,990 variants, 436,809 indels; AMR 7,290,008 variants, 493,933 indels; AFR 8,723,314 variants, 587,595 indels). Closely related subjects (plink pi_hat > 0.3) were excluded from the analysis.

#### 3: RNA-seq processing

##### 3.1: Alignment

Individual study RNA-seq data were acquired and uniformly processed. FastQC (v0.11.9) (*77*) was first run on FASTQ files for pre-alignment quality control. FASTQ files were then aligned to the hg19 reference genome with GENCODE v29lift37 (*78*) annotations using STAR-2.7.3a (*79*). Per-sample 2-pass mapping mode was used to improve novel junction discovery. Alignment quality control metrics were calculated using PicardTools (2.21.7) (<http://broadinstitute.github.io/picard/>) and compiled using MultiQC (v1.9ev0) (*80*).

##### 3.2: Sample swap check

Sample swaps and contamination were evaluated by calling SNPs from BAM alignments and merging with the imputed genotype. Plink --genome-full was used to calculate IBD/IBS sharing of all pairs of genotype ID (including both imputed and BAM-called). PI_HAT (proportion of IBD) and Z0 (P(IBD)=0) in the plink.genome output file were checked to determine whether there were any sample swaps. Imputed/BAM-called pairs from the same subject showed high levels of inbreeding (PI_HAT~1, Z0~0); while pairs from different subjects showed low levels of inbreeding (PI_HAT~0, Z0~1). Related subject pairs, which were removed in downstream analysis, showed high levels of IBD sharing. No unexpected sample swaps were detected.

##### 3.3: Gene and isoform quantification

Isoform-level quantifications were calculated using Salmon (v1.0.0)(*38*) and GENCODE v33lift37(*78*) annotations and a decoy sequence-aware index. generateDecoyTranscriptome.sh provided by Salmon was used to generate the index. Sequence bias and GC bias were corrected, and selective alignment of the sequences was enabled (--validateMappings --useEM --seqBias --gcBias). All subjects’ quantifications and gene-level quantifications were compiled using Tximport (1.14.0)(*81*). Raw counts, TPM, and length-scaled counts (countsFromAbundance="lengthScaledTPM") were generated for downstream analysis.

Genes/isoforms on chromosomes M, X, and Y were excluded for downstream analysis. Genes/isoforms with an expression level of TPM > 0.1 in more than 25% of subjects were included for downstream analysis. Counts were normalized and variances were stabilized using DESeq2::varianceStabilizingTransformation(*82*). Inter-subject connectivity was calculated using the R package WGCNA(*68*) and outlier subjects were excluded from the analysis (connectivity z-score < -3). Batch effects were corrected using sva::Combat(*83*). 31,947 genes and 127,986 isoforms (mapped to 31,121 genes) were included for QTL mapping.

##### 3.4: Splicing quantification

##### 3.4.1: Alignment

As above, FASTQ files were aligned to the hg19 reference genome using GENCODE v33lift37(*78*) annotation reference using STAR(*79*). To improve the sensitivity for discovering novel junctions, the multi-sample 2-pass mapping mode was used. At the end of the first pass of mapping, splice junctions (SJ) from all samples were filtered (excluding all MT junctions, all non-canonical SJ, SJ supported by multi-mappers-only, SJ supported by too few reads (<=2)). In the second pass, SJs from all samples were included in the mapping of all samples. WASP(*84*) filtering implemented in STAR was also applied in the second pass to reduce reference bias, and Samtools 1.9(*85*) was used to filter out reads that did not pass WASP (vW:i:[2-7]).

##### 3.4.2: Intron quantification and normalization

Introns were quantified using Leafcutter (v0.2.7) (*39*). Bam2junc.sh and leafcutter_cluster.py scripts provided by Leafcutter were used to quantify intron usage (excision ratio). Intron clusters were generated with the following options: >50 reads per cluster, >500kb intron length, >0.001 of reads in a cluster support an intron, >5 reads per intron (--minclureads 50 --maxintronlen 500000 --mincluratio 0.001 --minreads 5). 502,571 introns from 105,700 intron clusters and 22,751 corresponding genes were detected. The leafcutter script prepare_phenotype_table.py was then used to filter introns and prepare BED files. Non-autosomal introns, introns used in <40% of subjects, or with almost no variation were filtered out. Intron usage was standardized and quantile normalized. 273,167 introns from 50,311 clusters and 14,368 corresponding genes passed the filters and were included in sQTL mapping. ComBat was applied to the normalized data to remove batch effects.

##### 3.4.3: Intron annotation

We annotated the introns detected by Leafcutter in two slightly different ways. First, the script Gtf2leafcutter.pl was applied to the Gencode reference annotation GTF file to generate an annotation code: all_introns.bed.gz, threeprime.bed.gz, fiveprime.bed.gz, and all_exons.bed.gz. All detected introns were mapped to annotated introns, and intron clusters were mapped to the most representive gene of the introns in this cluster. Introns were annotated as follows

- “Unknown_strand”: intron is in a cluster with an unknown gene strand
- “Cryptic_unanchored”: intron is in a cluster with a known gene strand, but the intron does not have a known gene with 5’ or 3’ splice junctions
- “Cryptic_threeprime”: intron is in a cluster with + gene strand, known gene on 5’ end, unknown gene on 3’ end OR intron is in a cluster with - gene strand, known gene on 3’ end, unknown gene on 5’ end
- “Cryptic_fiveprime”: intron is in a cluster with + gene strand, known gene on 3’ end, unknown gene on 5’ end OR intron is in a cluster with - gene strand, known gene on 5’ end, unknown gene on 3’ end
- “Cryptic”: intron is in a cluster with unknown gene strand AND intron has known gene on 5’ OR 3’ end
- “Annotated”: intron has known gene on both ends
- “Novel annotated pair”: both are annotated but never in the same junctions

Second, we followed the approach taken by GTEx(*16*) to annotate introns by matching them with annotated exons. Briefly, each intron discovered by Leafcutter was matched with annotated exons by 3’ or 5’ junctions; then an intron cluster was mapped to all the genes with introns for which this cluster were matched. One cluster can map to multiple genes. In this way, 71,429 out of 105,700 clusters were mapped to genes; 5,132 clusters were mapped to more than 1 gene. 263,331 out of the 273,167 introns that we tested for sQTL have mapped genes in this way. Note that GTEx filters out introns that are not mapped to any genes; however, we only used this cluster-to-gene mapping file for grouped permutation-based sQTL identification (see Section 4.4).

#### 4: *cis*-xQTL mapping

##### 4.1: Covariate selection

Age and sex information from each of the donor samples were compiled from each study. Unavailable sexes were inferred from expression of *XIST* and aggregated expression of chrY genes. To account for unknown technical and biological factors, we employed the Hidden Covariates with Prior (HCP) (*86*) method using normalized expression values with a prior informed by PicardTools sequencing QC metrics. Age, sex, the top 5 genotype PCs, and HCP factors were included as covariates in QTL mapping.

##### 4.2: *cis*-eQTL mapping

*cis*-eQTL mapping was conducted using FastQTL v2.0(*87*). The *cis*-window was defined as 1 Mb up- and downstream around a gene’s transcription start site (TSS). Gene expression was standard normalized (--normal). To identify the optimal number of HCP covariates to include, a nominal association for each “gene-*cis* variant” pair was calculated using FastQTL in nominal mode, and multiple test-corrected with FDR < 0.05. The number of optimal HCPs was then selected by optimizing for the number of eGenes with nominally significant eQTLs. To discover eGenes (genes with at least one significant *cis*-eQTL), FastQTL adaptive permutations (--permute 1000 10000) were then run with the optimal number of HCP. Beta-approximated permutation p-value was multiple test-corrected using Storey-Tibshirani’s method(*40*), and eGenes were defined as those with a qvalue < 0.05. A nominal p-value threshold was calculated for each gene (following GTEx (*16*) and using qbeta to map the permutation adjusted p-value back to a nominal p-value threshold), and QTLtools v1.0(*41*) conditional mode was run to identify conditionally independent eQTL signals.

##### 4.3: *cis*-isoQTL mapping

*cis*-isoQTLs were mapped in a similar way as *cis*-eQTL. FastQTL nominal pass mode was first run with all expressed isoforms and variants within 1 Mb of the isoforms’ TSS. Then, using the optimized number of HCP factors as covariates, we first ran “naive” adaptive permutations (--permute 1000 10000), where isoforms are treated as independent from each other, and isoQTLs with a qvalue < 0.05 were identified as significant. Next, to account for the dependence between isoforms of a gene, we used grouped permutations as described in(*16*), where adaptive permutations were run jointly on all isoforms of a gene, and isoGenes were defined as those with a qvalue < 0.05.

##### 4.4: *cis*-sQTL mapping

HCP factors were generated using normalized intron usage values and a prior derived from PicardTools metrics. FastQTL nominal pass mode was run with all introns that passed filters, using variants within 1Mb of the intron body. The default 100kb window gave low p-values near the *cis* window boundaries, so we decided to use 1 Mb, as had been done for eQTLs and isoQTLs. Adaptive permutations (--permute 1000 10000) were run with the optimal number of HCP factors, and sQTLs with qvalue < 0.05 were identified as significant. Next, to account for the dependent structure of multiple introns and intron clusters of a gene, we ran grouped permutations, and sGenes were identified as those with a qvalue < 0.05.

#### 5: Inversion-associated eQTLs

Large-scale recurrent inversions were called in our fetal brain dataset using scoreInvHap (v1.10.0)(*88*). 17 common inversions had a sufficient number of surrounding SNPs in our QCd genotype data and could be imputed. A linear model was then fit to test for the association between each inversion and gene expression across the transcriptome. The same set of covariates was used as in *cis*-eQTL mapping. Specifically, covariates include age, sex, the top 5 genotype PCs, and 90 HCP factors. FDR correction was used to account for multiple testing within each inversion. Associations with FDR < 0.05 were reported as significant.

#### 6: Population-specific *cis*-xQTL mapping

To investigate population-specific genetic regulation, we separated genotype and gene/isoform/splicing data into the three major populations with N >100: EUR (N = 292), AMR (N = 164), and AFR (N = 145). Among genotyped variants, MAF and HWE filters and genotype PCs calculation were then performed separately in each population. We followed the *cis*-xQTL mapping approach for the multi-ethnic dataset and mapped *cis*-e/iso/sQTL in the three populations.

#### 7: cis-xQTL effect sizes

*cis*-xQTL effect sizes were measured by two metrics: allelic fold change (aFC) (*47*) and linear regression slope (beta). To compare *cis*-eQTL effect sizes between fetal and adult QTL reference panels, we calculated aFC in log2 scale for permutation-derived eGene-primary eQTL pairs in the fetal brain dataset, GTEx v8 Brain Cortex (*16*), and PsychENCODE (*13*).

#### 8: *cis*-xQTL overlap

We investigated the overlap between *cis*-e/iso/sQTLs using Storey’s pi1 statistic implemented in the qvalue R package(*40*). To estimate the proportion of eQTLs that are also truely associatiated with an isoQTL (pi1), we took the significant primary eGene-eQTL pairs passing the permutation threshold and matched them to all gene-SNP pairs’ association in isoQTL. We calculated pi1 as 1-(qvalue(pval_nominal_iso))$pi0. For eGene-eQTL pairs that are missing in isoQTL, we randomly assigned a nominal p-value sampled from a uniform null distribution. We did the same for all pairs between e/iso/sQTLs. For isoQTL and sQTL nominal associations, we first mapped isoforms and introns to genes as described in previous sections, and only kept gene-SNP pairs with the lowest nominal p-value.

#### 9: Functional enrichment of *cis*-xQTL

We generated genomic annotations using the following resources: Ensembl regulatory build (*89*) (homo_sapiens.GRCh37.Regulatory_Build.regulatory_features.20201218.gff.gz; annotations include: TF_binding_site, promoter_flanking_region, promoter, open_chromatin_region, enhancer, CTCF_binding_site), and Ensembl’s Variant Effect Predictor VEP v102 (*90*) on fetal brain genotype data (GENCODE v33lift37 annotation and genome fasta; using cache in homo_sapiens/102_GRCh37/ which has GENCODE v19, only annotations supported by GENCODE v33 was selected; annotation consequences include: synonymous_variant, stop_gained, splice_region_variant, splice_donor_variant, splice_acceptor_variant, non_coding_transcript_exon_variant, missense_variant, intron_variant, frameshift_variant, 5_prime_UTR_variant, 3_prime_UTR_variant). Functional enrichment analyses were performed using torus (*91*), with the genomic annotations, and all *cis* associations calculated by FastQTL (torus -d {fastqtl_allpairs} --fastqtl -est -annot {annot}).

#### 10: Gene constraint

To investigate the level of intolerance to loss-of-function mutation of genes, we downloaded ExAC data from gnomAD (*35*). We filtered for autosomal protein-coding genes and used the pLI score as a measure for gene constraint.

#### 11: Fine-mapping of *cis*-xQTL

We performed QTL fine-mapping using the Sum of Single Effects Regression (SuSiE) as implemented in the susieR_0.11.42 package (*42*). The fine-mapping was performed for each eGene, isoform with permutation-significant isoQTL, and intron with permutation-significant sQTL and variants within 1 Mb from the feature. The genotype VCF file used for QTL mapping was converted to a dosage matrix using bcftools 1.9 (*92*). Genotype dosage and gene/isoform expression or intron splicing files were residualised by the same set of covariates as those used in the FastQTL permutations. Fine-mapping was performed with the following parameters: L = 10, estimate_residual_variance = TRUE, estimate_prior_variance = TRUE, scaled_prior_variance = 0.1, compute_univariate_zscore = TRUE, min_abs_corr = 0. We then constructed credible sets (CS) for the features by extracting variants with posterior inclusion probabilities (PIPs) summing up to 95%. As described in the SuSiE paper, we used CS purity as a measure of the correlation of variants in that CS and filtered for high-purity CS for downstream analysis.

For population-specific *cis*-eQTLs, we performed SuSiE fine-mapping separately within each population. For the multi-ethnic dataset, we first performed SuSiE fine-mapping, and then to leverage population-specific linkage disequilibrium (LD), we performed multi-ethnic fine-mapping using PAINTOR (*48*, *49*), which uses genomic annotations (see Section 9) and population-specific summary statistics and genetic backgrounds. For the 982 permutation eGenes shared between EUR, AMR, and AFR, we ran PAINTOR with the *cis* variants shared by the three populations. We enumerated with 2 casuals at maximum (-enumerate 2) and constructed 95% CS based on PIPs.

#### 12: Age specificity of genetic regulation

We stratified the EUR samples into first and second trimesters, and mapped trimester-specific *cis*-e/iso/sQTL (eQTL Tri1 N = 137, Tri2 N = 142; isoQTL Tri1 N = 136, Tri2 N = 139; sQTL Tri1 N = 141, Tri2 N = 143. The sample sizes slightly differ because gene and isoform expression outlier samples were excluded from the analysis, see Section 3.3). We used the same set of covariates as described in Section 4.1.

We estimated heritability for both gene expression and splicing phenotypes in European individuals by fitting a univariate variance component linear mixed model via the restricted maximum likelihood (REML) approach using the majorization-minimization (MM) algorithm implemented in the MultiResponseVarianceComponentModels.jl (*93*). We specified three variance components corresponding to genetic effects from *cis*-SNPs (defined as SNPs within 1 Mb around gene TSS and intron body), genetic effects from trans-SNPs (defined as SNPs that do not fall within the previously described *cis*-window), and residual effects. For each gene/intron, we first regressed out fixed effects covariates from the complete EUR expression files, constructed two separate classic genetic relationship matrices (GRMs) for *cis*-SNPs and trans-SNPs using SnpArrays.jl (*94*), and then estimated variance components for each trimester separately. Additionally, a sliding window of samples by age was analyzed—that is, after sorting the samples by age, variance components were estimated for batches of 150 samples with the window shifting 25 samples by batch. Heritability estimates and their standard errors were subsequently calculated using the delta method, which is built into the Julia package.

#### 13: GWAS summary statistics

To investigate the overlap between fetal brain xQTLs and neuropsychiatric disorder GWAS results, we downloaded GWAS summary statistics for the following traits: schizophrenia (SCZ) (*1*) (we used the European summary statistics), autism spectrum disorder (ASD) (*2*), bipolar disorder (BIP) (*56*), attention-deficit/hyperactivity disorder (ADHD) (*54*), major depression (MDD) (*95*). GWAS summary statistics were munged using munge_sumstats.py script from LD Score Regression (LDSC) (*6*, *50*).

#### 14: MESC

We applied Mediated Expression Score Regression (MESC) (*53*) to estimate the proportion of neuropsychiatric disease heritability mediated by genes, isoforms, and introns, respectively. QTL effect sizes were estimated using individual-level expression and genotype data to reduce noise (--compute-expscore-indiv). LD scores and LD weights calculated from the 1000 Genomes Phase 3 EUR samples (*44*) were included in the analysis. By default, LD scores are computed from 1000 Genomes Phase 3 stratified over a modified version of the baselineLD model v2.0 (<https://data.broadinstitute.org/alkesgroup/LDSCORE/>).

#### 15: Stratified LD score regression

To investigate the enrichment of neuropsychiatric GWAS heritability among fetal brain QTLs, we first generated the maxCPP annotation(*51*) for the following QTL datasets: *cis*-eQTLs, *cis*-isoQTLs, *cis*-sQTLs, and trimester-specific e/iso/sQTLs. To construct this probabilistic maxCPP annotation, we first ran SuSiE fine-mapping for each dataset(*42*). For each variant in the genome, we then assigned an annotation value based on the maximum fine-mapping PIP across all molecular features within which the variant lies in the SuSiE 95% CS. Variants that do not belong to any of the SuSiE CSs were assigned an annotation value of 0. We restricted to 1000 Genomes EUR HapMap3 variants for the analysis. The corresponding maxCPP annotation for GTEx v8 Brain Cortex was also downloaded (*16*, *51*).

Next, we applied stratified LD score regression (S-LDSC) (*6*) using the maxCPP annotations. S-LDSC was performed using the 1000 Genomes EUR reference panel (*44*), restricted to HapMap3 SNPs, and corresponding regression weights. We applied S-LDSC for each GWAS and QTL dataset pair, jointly with 53 baseline annotations (<https://alkesgroup.broadinstitute.org/LDSCORE/>). We used two metrics to measure the importance of an annotation: the enrichment and effect size z-score. The enrichment of an annotation is defined as the proportion of heritability captured by the annotation divided by the proportion of SNPs in that annotation (Pr(h2)/Pr(SNP)). The effect size z-score (Coefficient_z.score = Coefficient/Coefficient_std_error) is an estimate of how much more significantly the annotation explains the GWAS statistics on top of the other annotations in the model.

#### 16: Isoform-level TWAS

To identify genes and their isoforms whose *cis*-regulated expression is associated with neuropsychiatric diseases, we performed an isoform-level transcriptome-wide association study (isoTWAS) (*65*). We generated SNP weights using our uniformly processed fetal brain dataset (full dataset N = 629). We used the imputed genotype data restricted to HapMap 3 variants only and expression corrected by the same set of covariates as in *cis*-isoQTL mapping (age, sex, 5 genotype PCs, and HCPs). Briefly, we built expression models using multivariate predictive methods that model all heritable isoforms of a gene jointly using either (1) multivariate elastic net, (2) multivariate SuSiE regression, or (3) multivariate LASSO regression with simultaneous covariance estimation using graphical LASSO. These multivariate methods account for the correlation between isoforms of the same gene to increase the prediction accuracy of each individual isoform, compared to methods that consider each isoform separately. Optimal model weights using all SNPs within a 1Mb window of the genes were trained by selecting the model with the best 5-fold cross-validation R^2^. We filtered for isoforms for cross-validation R^2^ >0. A stepwise trait mapping procedure is used in isoTWAS (*65*). First, isoform associations from the same gene family to the gene-level using the aggregated Cauchy association test (*96*), and gene-level P-values are adjusted for multiple testing burden using a Bonferroni correction. Second, for isoforms of a gene that passes Bonferroni correction, the family-wise error rate for isoforms of the same gene is controlled using Shaffer’s modified sequentially rejective Bonferroni correction (*97*). Through this process, we identified both gene-disease and isoform-disease associations with the LD structure calculated from our data and schizophrenia GWAS summary statistics. We then prioritized isoform-disease associations using a permutation-based test (1000 permutations of the SNP-isoform weights). This permutation test assesses whether the SNP-isoform weights add any mechanistic insight beyond the SNP-disease associations from the GWAS.

#### 17: FOCUS

To leverage local LD and prioritize candidate isoforms for isoTWAS analysis, we performed probabilistic fine-mapping using FOCUS (*98*). FOCUS models the correlation among isoform-specific signals and estimates the posterior probability of an isoform explaining the observed isoTWAS association signal in the risk region. We generated 90% credible sets of causal isoforms for any set of isoforms that showed isoTWAS associations.

#### 18: eCAVIAR

eCAVIAR (v2.2) (*9*) was used to prioritize likely causal variants responsible for both QTLs and GWAS. For each GWAS described in Section 13, we extracted variants within 1 Mb around the index variant for each GWAS loci. For each locus, summary statistics for variants that are also in the 1000 Genomes EUR reference panel (*44*) were extracted. Candidate target features (i.e. genes, isoforms, introns) were defined as features with at least one of the GWAS locus variants as nominal QTL (FDR < 0.05). Next, for each candidate target feature of the locus, we extracted the overlapping variants between GWAS and the feature’s *cis* variants. We calculated the LD of these variants for GWAS using 1000 Genomes EUR reference panel and for the multi-ethnic fetal brain samples using in-sample genotypes. Finally, we ran eCAVIAR and calculated the colocalization posterior probability (CLPP) for each variant (a maximum of two possible causal variants for each locus was assumed: eCAVIAR -f 1 -c 2 -r 0.95). Significant colocalization was defined as CLPP > 0.01. Sashimi.py (*99*) was used to create the SP4 read density plot and GeneticsMakie.jl (*93*) was used to create the LocusZoom plots.

#### 19: MRLocus

To estimate gene-to-trait effect sizes, MRLocus-eCAVIAR (*67*) was run on conditional fetal brain eQTL (from this study) and Schizophrenia GWAS (*1*) summary statistics (European ancestries; 53,386 cases and 77,258 controls; <https://figshare.com/articles/dataset/scz2022/19426775>). Gene-to-trait effect sizes were only tested within a subset of genes already shown to be associated with schizophrenia risk via TWAS. Among 199 genes discovered through TWAS, 14 genes were selected with 3 or more instruments: conditionally distinct sets of SNPs associated with those eGenes. For each signal cluster (a set of SNPs associated with gene expression, which is conditionally distinct and LD-independent from other sets), we extracted beta and standard error (SE) values from both eQTL and GWAS summary statistics. Since eQTL and GWAS were conducted within different populations, we prepared two LD matrices (*r*) based on (1) individuals enrolled in the eQTL population composed of multiple ancestries and (2) pre-calculated non-Finnish European LD matrices provided by the gnomAD v2 (*100*) for the GWAS population. Only SNPs that existed in all datasets (i.e. eQTL, GWAS and gnomAD reference panel) were used for further analysis. Running MRLocus-eCAVIAR entailed using eCAVIAR (v2.2) (*9*) to estimate posterior probabilities of colocalization within each signal cluster to choose candidate SNPs, followed by running MRLocus (v0.0.24) gene-to-trait slope estimation over signal clusters. For signal clusters that straddled a 500kb boundary from the gene TSS, SNPs that were greater than 500kb away from TSS were filtered from the cluster before running eCAVIAR. To reduce dependency across clusters, a cluster was trimmed and not used in slope estimation if the eQTL SNP (instrument) had r^2^ > 0.05 to any other cluster's eQTL candidate SNP, following the trimming procedure described in (*67*). Per locus p-values testing the null hypothesis of no gene-to-trait effect were corrected for multiple testing using the Benjamini-Hochberg method, providing FDR control at 20% for raw p-values less than 0.078.

#### 20: WGCNA

##### 20.1: Preparation of Count Data

We performed robust Weighted Gene Correlation Network Analysis (WGCNA) on gene-level and isoform-level quantifications in order to group fetal genes into disease-relevant networks. First, unscaled and untransformed gene- and isoform-level counts for all the samples were obtained following the procedure outlined in 3-3. Outlier sample removal and expression filtering resulted in a gene-level count matrix of 31,947 genes and 642 samples and an isoform-level count matrix of 127,986 transcripts and 639 samples. Next, a vector of each sample’s study of origin was fed to ComBat-seq in the *sva* R package (version 3.35.2) to generate batch-corrected count data. CTF library normalization (*101*) was performed on the batch-corrected count data using TMM normalization factors obtained from the *edgeR* R package (version 3.36.0). Briefly, a CTF factor for each sample was generated by multiplying each sample’s edgeR-derived normalization factor by the sample’s library size. Then, a scaled CTF factor was obtained by dividing each CTF factor by the overall smallest CTF factor. Finally, each sample’s expression was divided by the scaled CTF factor, incremented by 1, and log2 transformed, resulting in a pseudo-count matrix of normalized expression. The gene-level samples (n = 642) were subsequently split by trimester into tr1-gene (n = 212) and tr2-gene (n = 427), and by sex into xx-gene (n = 302) and xy-gene (n = 340).

##### 20.2: Network Generation

Gene- and isoform-level signed networks were generated with the WGCNA R package (version 1.69.0). First, for each log2-normalized expression matrix, a soft-threshold power was selected to minimize mean connectivity and maintain a scale free R^2^ > 0.8. For each gene-level expression matrix, a topological overlap matrix (TOM) was robustly generated using the consensusTOM function on a vector of 100 downsampled pseudo-count matrices, each containing ⅔ of the respective samples. This robust WGCNA approach (rWGCNA) reduces the likelihood that resulting modules will capture signals corresponding to sample subpopulations instead of sample-wide biological signals (*68*). For the isoform-level expression matrix, since rWGCNA was too computationally expensive, a vector containing the single original pseudo-count matrix was processed through the consensusTOM function. Network dendrograms for each TOM were generated using average linkage hierarchical clustering of the dissimilarity TOM (1 - TOM). Modules were subsequently constructed using the cutreeHybrid function with a minimum module size of 200, deep split parameter of 4, pamStage of 1, pamRespectsDendro of 0, and cut height of 0.999. Highly correlated modules were combined with the mergeCloseModules function using a merge cut height of 0.225.

##### 20.3: Cell Type Enrichment

Within each module, genes with a kME > 0.5 were compared against known fetal brain cell markers(*36*) using over-representation analysis. The original 16 cell types were collapsed into 9 broad cell classes, corresponding to hierarchical clustering of these developmental cell populations(*59*). Fisher's exact test p-values for gene-marker overlap were collected into a table and subsequently FDR-corrected to control for multiple comparisons. Module parameters were selected to maximize cell type enrichment.

##### 20.4: Disease Enrichment

We computed each module’s enrichment for common and rare disease variants to identify key disease-associated developmental modules.

For common variation, we utilized summary statistics from the largest available GWAS across major neurodevelopmental disorders (Section 13). Each module’s SNP-level disease heritability was estimated with S-LDSC. First, module genes were assembled into lists, and genomic annotations were generated using a 10kb window with GENCODE v33lift37 as a reference. Then, LD scores were calculated for each set of annotation files, and partitioned heritability was used to estimate the SNP heritability for each module was jointly compared with a baseline set of 53 genomic annotations. Finally, each module’s SNP heritability was divided by the proportion of SNPs annotated within that module. Significance was assessed by converting the regression coefficient (tau) z-scores into p-values. Additionally, gene-level disease heritability for each module was estimated using MAGMA(*102*). For each GWAS, SNP-level effect sizes were converted to gene-level effect sizes using the default MAGMA annotation set(*103*). For each network, a two-column file containing module membership status and gene identity was generated and run through MAGMA in gene-set analysis mode. The resulting module enrichment p-values were subsequently FDR-corrected for multiple comparisons.

For rare variation, we used compiled curated gene lists for each disorder: SFARI gene list for ASD(*104*), Epi25 for Epilepsy(*105*), BipEx for BIP(*106*), The Deciphering Developmental Disorders Study (DDD)(*107*), SCHEMA for SCZ(*108*). In addition, we downloaded TADA-based summary statistics for ASD and DDD from(*59*) and curated genes for epilepsy, as published by Polioudakis et al(*36*). A binary vector representing each gene’s membership with the module (with 1 for genes in the specified module and 0 otherwise) served as the dependent variable. For gene lists without summary statistics, a binary vector representing each gene’s membership in the curated gene list was used as the primary independent variable, and gene length, log10 of gene length, transcript length, and log10 of transcript length were used as covariates. For gene lists with reported p-values, a continuous vector containing -log10(p-value) for each gene measured by the study served as the primary independent variable, controlling for gene and transcript length as above. Logistic regression was then used to test for association, and the resulting odds ratios and p-values were collected into a table. All p-values were FDR-corrected to control for multiple comparisons.

##### 20.5: Biological Pathway Enrichment

To place modules into biological context, GO term enrichment was performed using the gprofiler2 R package (version 0.2.1). For each module, genes were assembled into a vector, arranged from highest to lowest kME, and processed through the gost function as an ordered query. In addition, a vector consisting of all expressed genes in our fetal dataset was passed as a custom background to the gost function. The resulting GO enrichment table was filtered for significant results and term sizes smaller than 2,000, and plotted using the gostplot function.

##### 20.6: Module interaction eQTL mapping and GWAS colocalization

To identify *cis*-eQTLs conditional on the module enrichment, we tested the interaction between genotypes and modules using a linear regression model. The formula is exp~g+m+g*m+c, where exp is the gene expression, g is the genotype, m is module eigengenes which is the first principal component of expressions of module member genes, and c is the covariate matrix that was used in bulk tissue eQTL mapping. The covariates include sex, age, genotype PCs, and HCP factors. The correlations between HCPs and module eigengenes are lower than 0.5, suggesting the HCP factors are robust to modules. TensorQTL (*109*) was used to compute the significance of the interaction term. Variants within the 1Mb window of TSS of each tested gene were included. Variants with MAF < 0.05 were removed. The phenotype input is the same matrix used in bulk tissue eQTL mapping. The top SNP with the most significant p-value for each gene was selected. P values were firstly corrected at the gene level using eigenMT. Then the genome-wide significance was computed by the Benjamini-Hochberg method. Next, we followed the eCAVIAR pipeline as described in Section 18, and ran colocalization analysis on 120 SCZ locus-isomodule-gene, and 3220 SCZ locus-genemod-gene tuples. Variant with CLPP > 0.01 was identified as significant colocalization.

To test the cell-type-specificity of module eQTLs, we used eQTLs from cultured neurons and progenitor cells as a reference (*72*). We calculated the pi1 statistic to quantify the sharing between the module and cell-type eQTLs using the qvalue package (*40*). The nominal p values of shared eSNP-eGene pairs in reference data were used to calculate pi0 statistic. The pi1 was calculated as 1-pi0.

#### 21: CellWalker

We used the network-based CellWalker (*71*) method to map eQTLs to specific cell types through integration with fetal single-cell chromatin accessibility data (scATAC-seq). We built and tuned a network model using CellWalkR version 0.99 (*110*) with labels derived from Polioudakis et al. (*36*) and scATAC-seq data from Ziffra et al. (*111*). Label edges were generated using the mapSnapATACToGenes function of CellWalkerR with the gene activity matrix from Ziffra et al. and log-fold change expression in marker genes from Polioudakis et al. Cell edges were computed as the Jaccard similarity of each cell's genome-wide accessibility. We selected a label edge weight parameter of 100 using the tuneEdgeWeights function with the steps parameter set to three. Once the network model was built, we used the CellWalkR labelBulk function to score all bulk and cell-type specific eQTLs for all cell types in Polioudakis et al. We called an eQTL as being specific to a cell type if it received a label score greater than two for that cell type. We then extended this scoring scheme to generate multi-level cell types by averaging the label scores of each eQTL at each branch of the hierarchy of cell types as built using the CellWalkR clusterLabels function with the "maximum" option for the distance method, and scaled by 1.5 at each level. A count of cell-type labels for the outermost nodes and how many of the multi-level label scores were greater than two for the inner nodes was used to generate a hierarchical tree of enriched cell types.

### Supplementary Figures

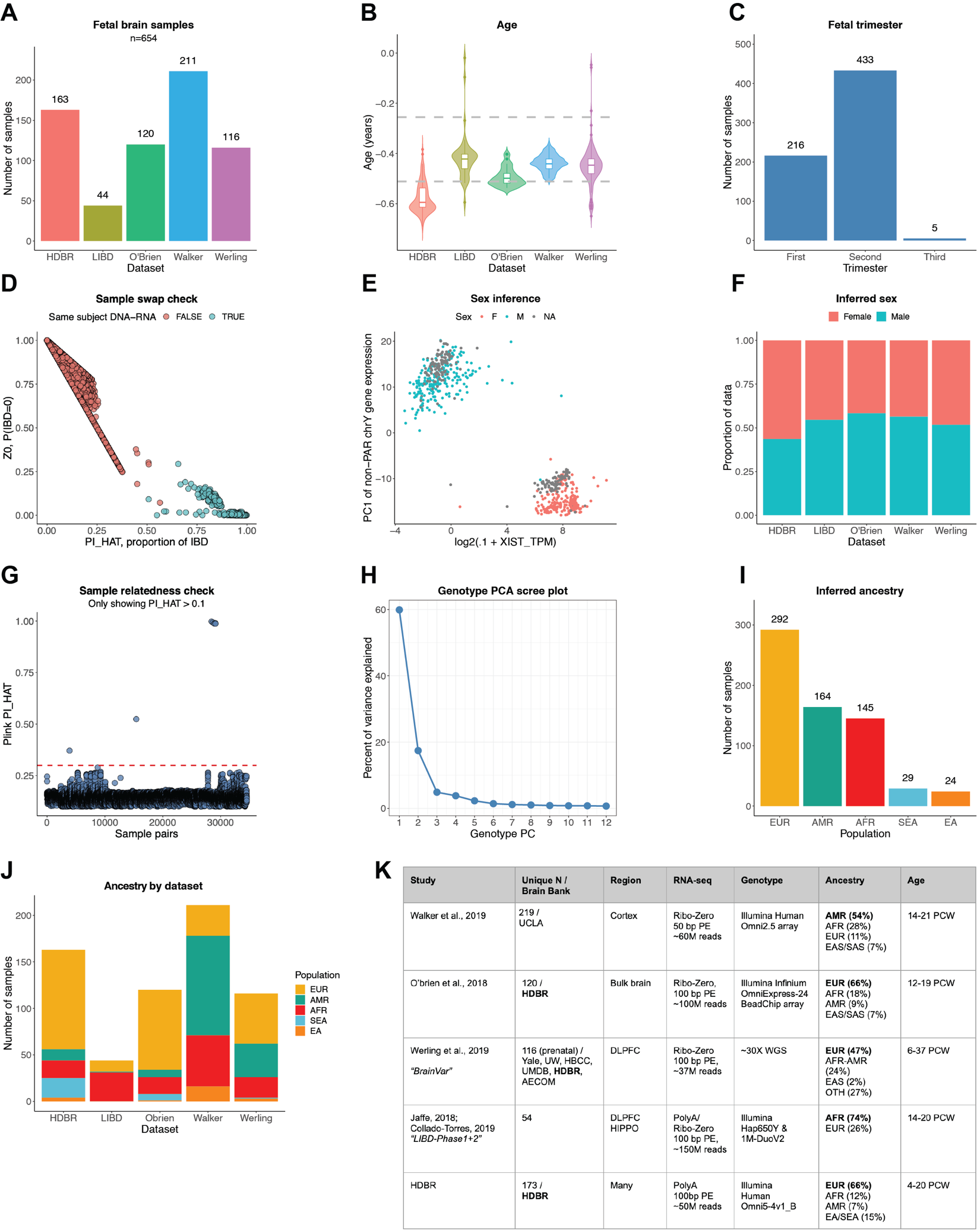

##### Fig. S1. Compilation of developmental brain datasets.

**(A)** Sample sizes of fetal brain studies. **(B)** Age of fetal brain samples, spanning from 4 to 39 PCW. **(C)** Trimester of fetal brain samples (Tri1: 4-13 PCW; Tri2: 14-26 PCW; Tri3: 27-39 PCW). **(D)** Sample swap check. **(E)** *XIST* gene expression versus PC1 of chromosome Y gene expression, colored by sex of fetal brain samples. Unknown sexes are inferred. **(F)** Inferred sex of fetal brain samples by study. **(G)** Sample relatedness check, as measured by PI_HAT. PI_HAT>0.3 samples were excluded from the analysis. **(H)** Scree plot of population PCA. **(I)** Number of fetal brain samples inferred as European (EUR), American (AMR), African (AFR), Southeast Asian (SEA), and East Asian (EA). **(J)** Inferred ancestry of fetal brain samples by study. **(K**) Overview of fetal brain datasets. 654 samples with high-quality matching DNA-RNA data were kept in the analysis.

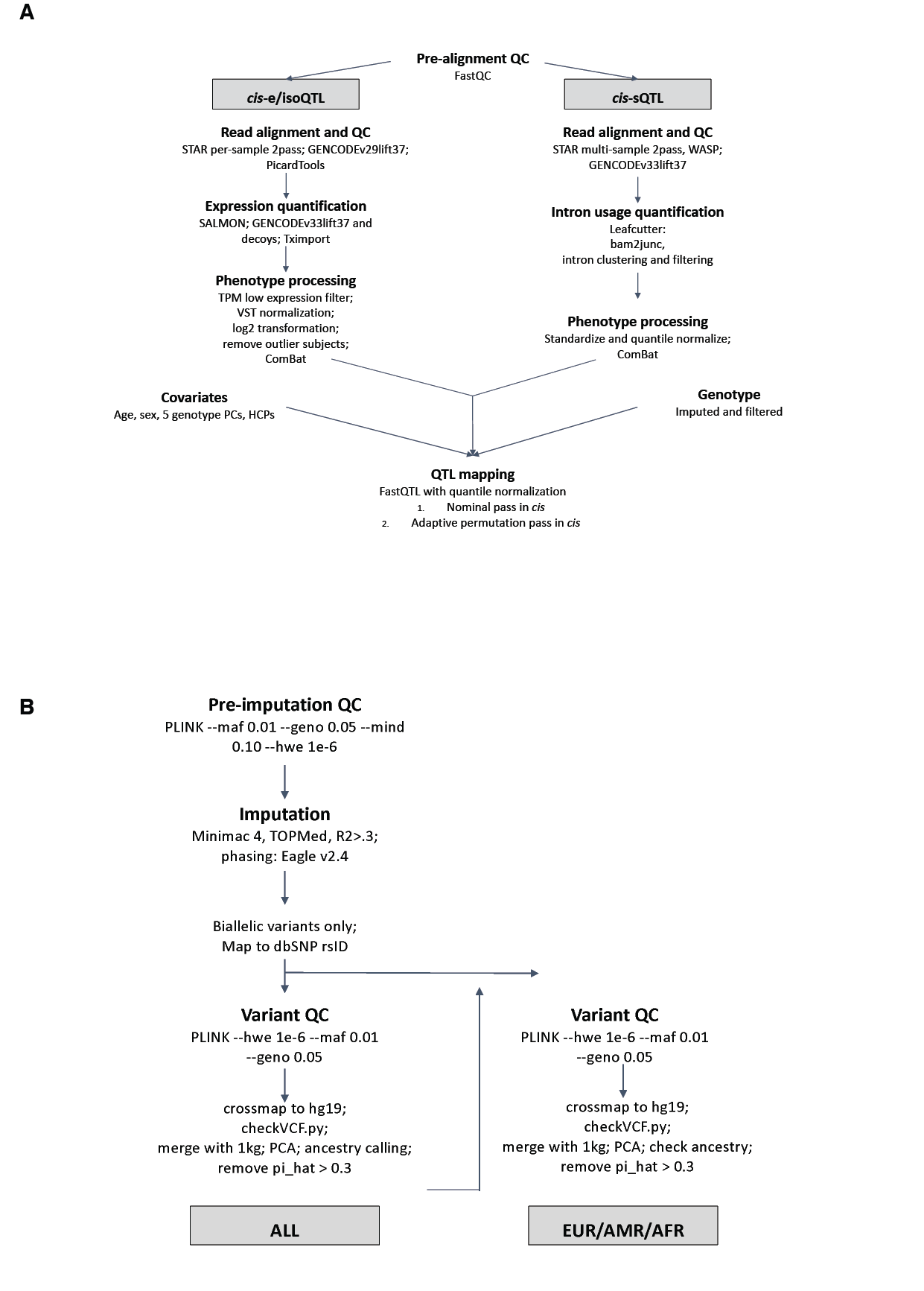

##### Fig. S2. Data analysis and integration pipeline.

**(A)** Analysis pipeline through which RNA-seq data of all samples were uniformly processed. **(B)** Analysis pipeline through which genotype data of all samples were uniformly processed.

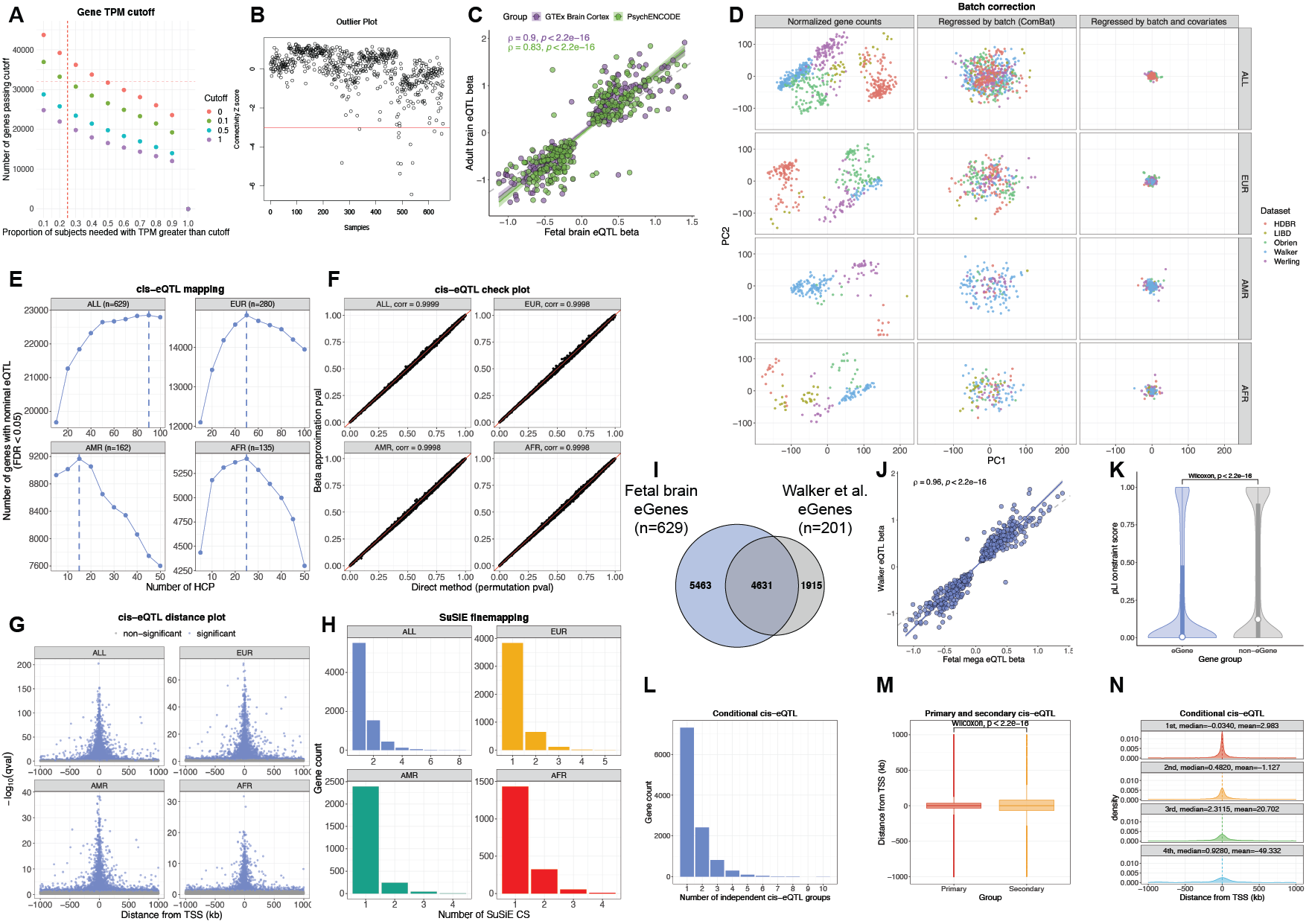

##### Fig. S3: *cis*-eQTL mapping and fine-mapping.

**(A)** Gene expression filtering. 31,947 genes with greater than 0.1 TPM in at least 25% of the samples were included in the analysis. **(B)** Gene expression outlier sample detection. Samples with connectivity Z score < -3 were excluded from the analysis. **(C)** Correlation of *cis*-eQTL effect size, measured by linear regression slope (beta), between fetal and adult brain. Each dot is a shared pair of eGene-primary eQTL between fetal brain and GTEx (247 pairs) or PsychENCODE (253 pairs). **(D)** Batch correction of gene expression. The top 2 principal components (PCs) were shown for before batch correction (left), after batch correction (middle), and after covariates correction (right). **(E)** Nominal *cis*-eQTL mapping. The optimal number of Hidden Components with Priors (HCPs) was determined by optimizing for the number of significant nominal eGenes. 90, 50, 15, and 25 HCPs were included in a permutation-based analysis for the multi-ancestry (ALL, n=629), EUR (n=280), AMR (n=162), and AFR (n=135), respectively. **(F)** Check plot of FastQTL beta approximation of permutation p value. **(G)** Distance from gene TSS of primary *cis*-eQTLs. **(H)** Number of SuSiE fine-mapping credible sets of eGenes in ALL, EUR, AMR, and AFR. **(I)** Comparison of eGenes between fetal mega-analysis dataset and Walker et al. **(J)** Correlation of *cis*-eQTL effect size measure by beta between fetal and Walker datasets. **(K)** pLI score of eGenes and non-eGenes in the fetal dataset. **(L)** Number of conditionally independent *cis*-eQTL groups. **(M)** Comparison of distances from gene TSSs between primary and secondary *cis*-eQTLs. **(N)** Comparison of distances from gene TSSs between the 1st to 4th ranks of *cis*-eQTLs.

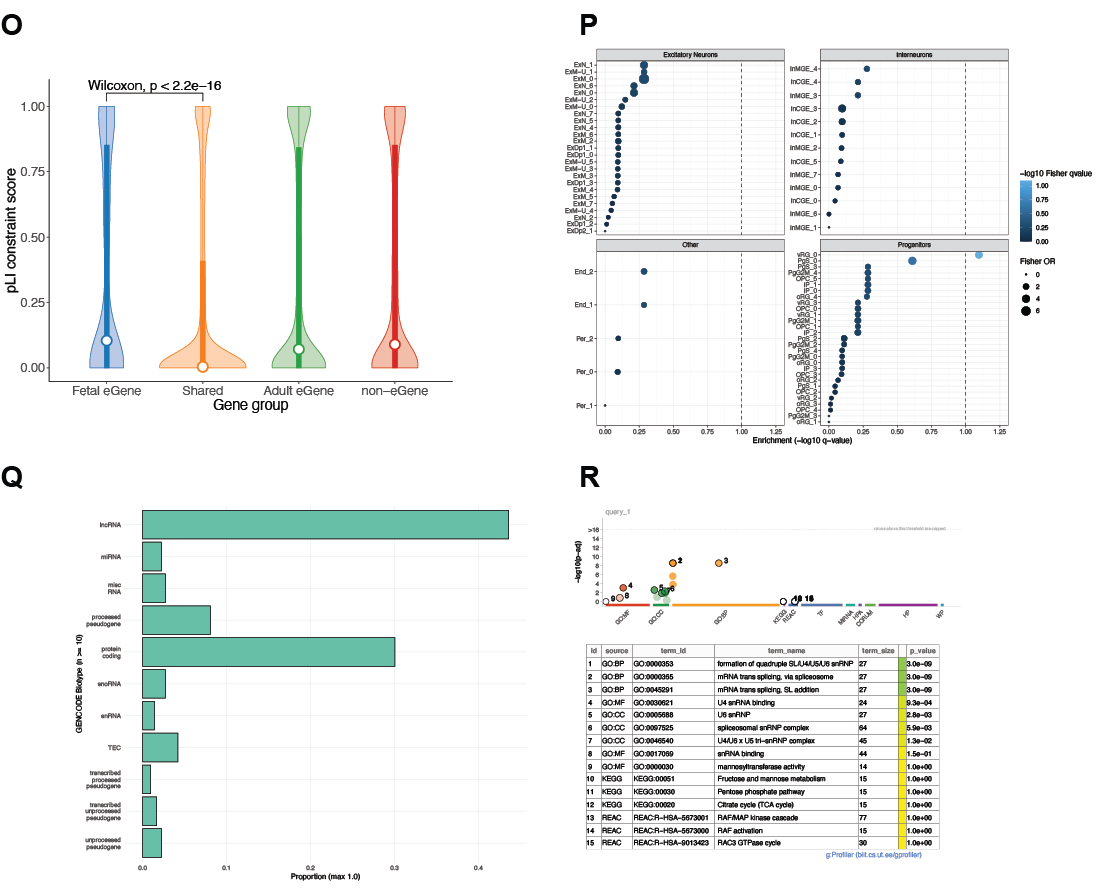

##### Fig. S3-continued: Characteristics of 2,488 fetal-specific eGenes.

(**O**) Tolerance to loss-of-function mutations, as measured by pLI score, of fetal-specific eGenes, adult-specific eGenes, shared eGenes, and non-eGenes. (**P**) Cell type enrichment of fetal-specific eGenes. (**Q**) Proportion of gene types of fetal-specific eGenes. (**R**) GO pathway enrichment of fetal-specific eGenes.

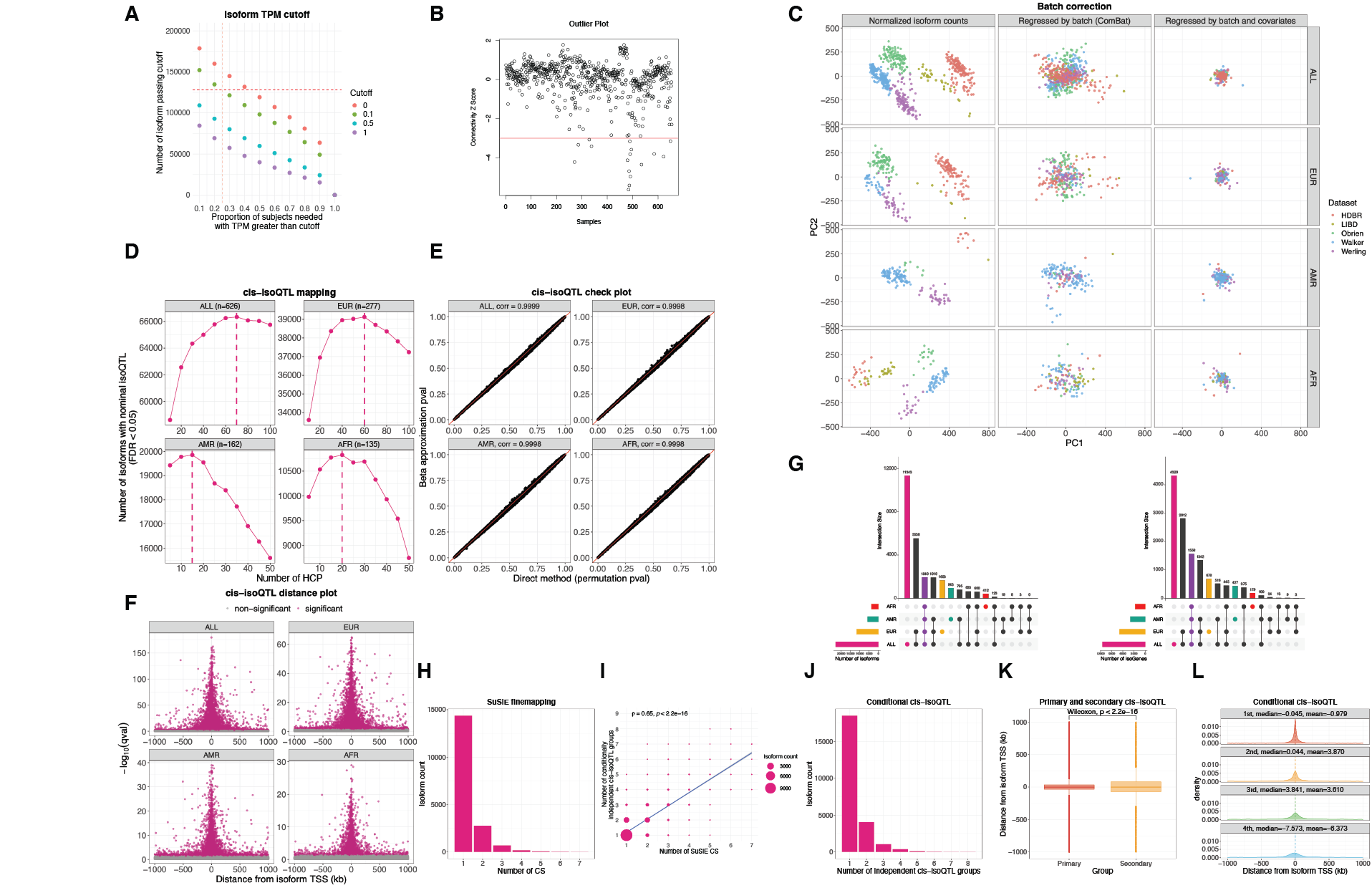

##### Fig. S4. *cis*-isoQTL mapping and fine-mapping.

**(A)** Isoform expression filtering. 127,987 isoforms with greater than 0.1 TPM in at least 25% of the samples were included in the analysis. **(B)** Isoform expression outlier sample detection. Samples with connectivity Z score < -3 were excluded from the analysis. **(C)** Batch correction of isoform expression. The top 2 principal components (PCs) were shown for before batch correction (left), after batch correction (middle), and after covariates correction (right). **(D)** *cis*-isoQTL nominal mapping. The optimal number of Hidden Components with Priors (HCPs) was determined as when the number of isoforms with nominal *cis*-isoQTLs was maximized. 70, 60, 15, and 20 HCPs were included in a permutation-bsed analysis for the multi-ancestry (ALL, n=626), EUR (n=277), AMR (n=162), and AFR (n=135) datasets, respectively. **(E)** Check plot of FastQTL beta approximation of permutation p value. **(F)** Distances from isoform TSSs of primary *cis*-isoQTLs. **(G)** Comparison of isoforms (left) and genes (right) identified with *cis*-isoQTLs discovered in ALL, EUR, AMR, and AFR. **(H)** Number of SuSiE fine-mapping credible sets of isoforms. **(I)** Number of fine-mapping credible sets versus number of conditionally independent *cis*-isoQTLs discovered. The size of the dots is scaled to designate the number of isoforms. **(J)** Number of conditionally independent *cis*-isoQTL groups. **(K)** Comparison of distances from isoform TSSs between primary and secondary *cis*-isoQTLs. **(L)** Comparison of distances from isoform TSSs between the 1st to 4th ranks of *cis*-isoQTLs.

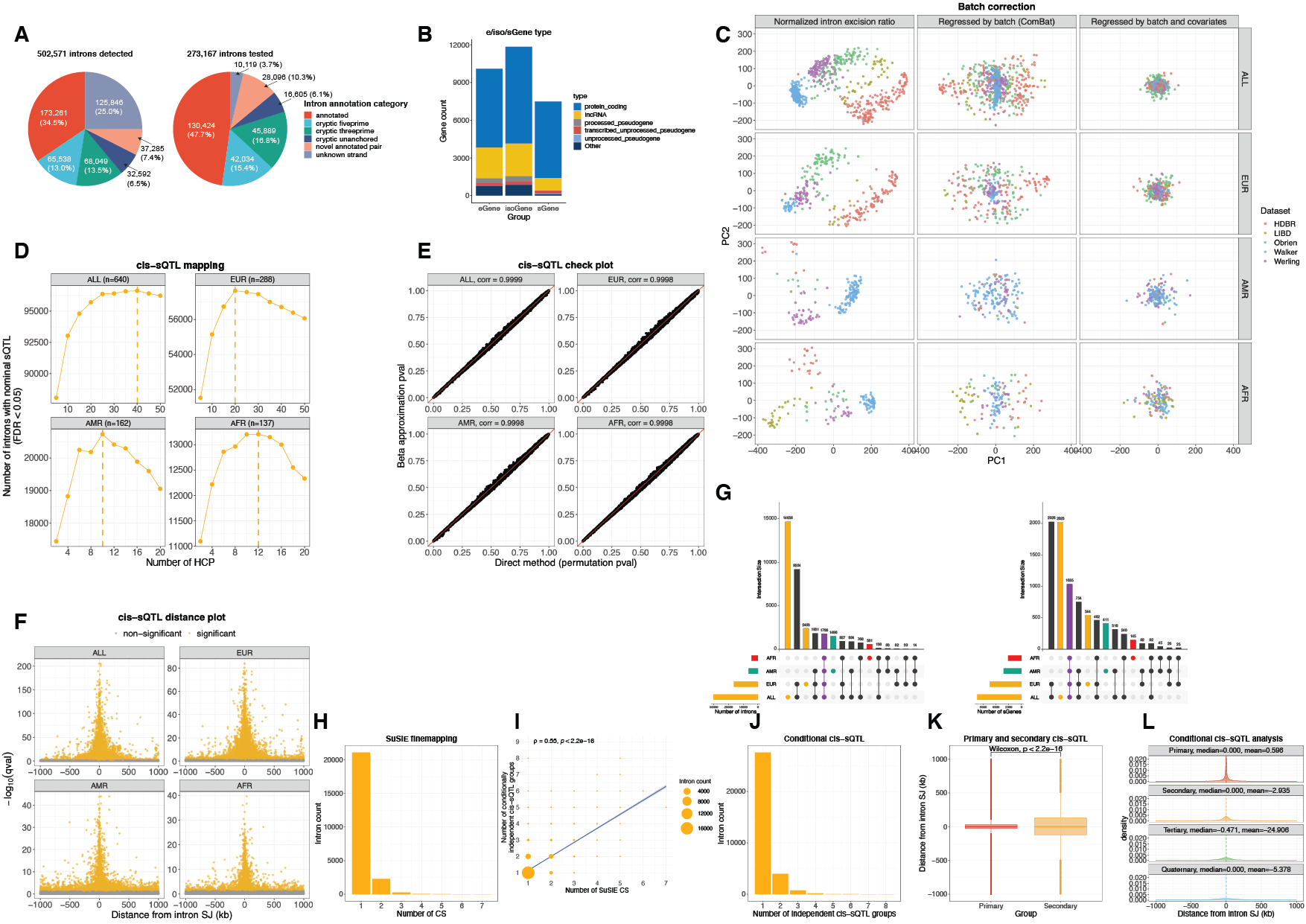

##### Fig. S5. *cis*-sQTL mapping and fine-mapping.

(**A**) GENCODE (v33) annotations of the 502,571 introns detected by Leafcutter (left), and the 273,167 introns that passed filters and were used to map sQTL (right). (**B**) Gene types of *cis*-eGenes, isoGenes, and sGenes. (**C**) Batch correction of intron quantification. The top 2 principle components (PCs) were shown for before batch correction (left), after batch correction (middle), and after covariates correction (right). (**D**) *cis*-sQTL nominal mapping. The optimal number of Hidden Components with Priors (HCPs) was determined as when the number of introns with nominal *cis*-sQTLs was maximized. 40, 20, 10, and 12 HCPs were included in permutation analysis for the multi-ancestry (ALL, n=640), EUR (n=288), AMR (n=162), and AFR (n=137) datasets, respectively. (**E**) Check plot of FastQTL beta approximation of permutation p value. (**F**) Distances from intron of primary *cis*-sQTLs. (**G**) Comparison of introns (left) and genes (right) identified with *cis*-sQTLs discovered in ALL, EUR, AMR, and AFR. (**H**) Number of SuSiE fine-mapping credible sets of introns. (**I**) Number of fine-mapping credible sets versus number of conditionally independent *cis*-sQTLs discovered. The size of the dots is scaled to designate the number of introns. (**J**) Number of conditionally independent *cis*-sQTL groups. (**K**) Comparison of distances from intron between primary and secondary *cis*-sQTLs. (**L**) Comparison of distances from introns between the 1st to 4th ranks of *cis*-sQTLs.

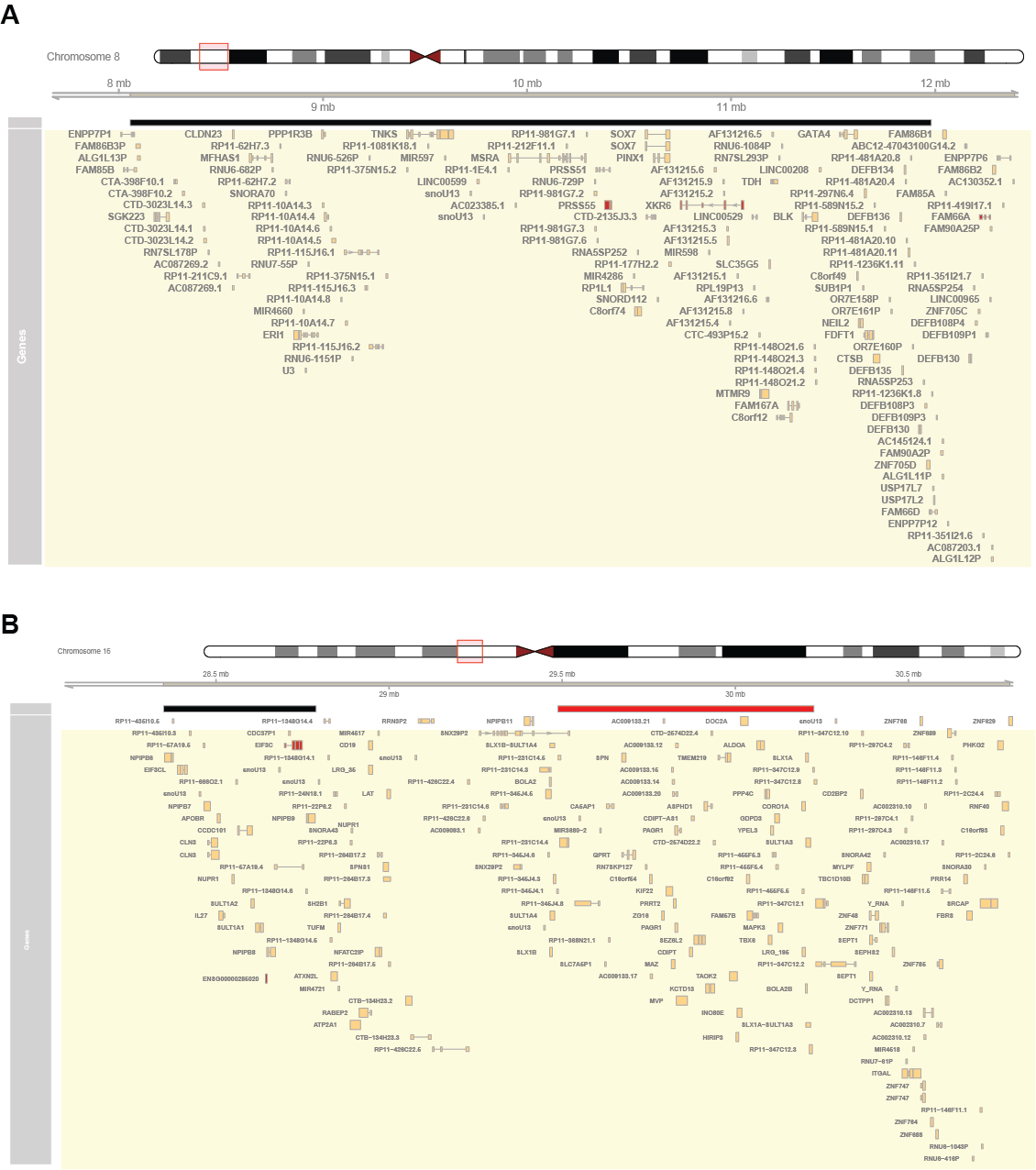

##### Fig. S6: Inversion eQTL.

Genomic location of inversion (black bar) and their eGenes *in cis* (red colored genes) for regions at **(A)** 8p23.1 and **(B)** 16p11.2. At 16p11.2 a rare CNV region near the inversion is highlighted with red bar.

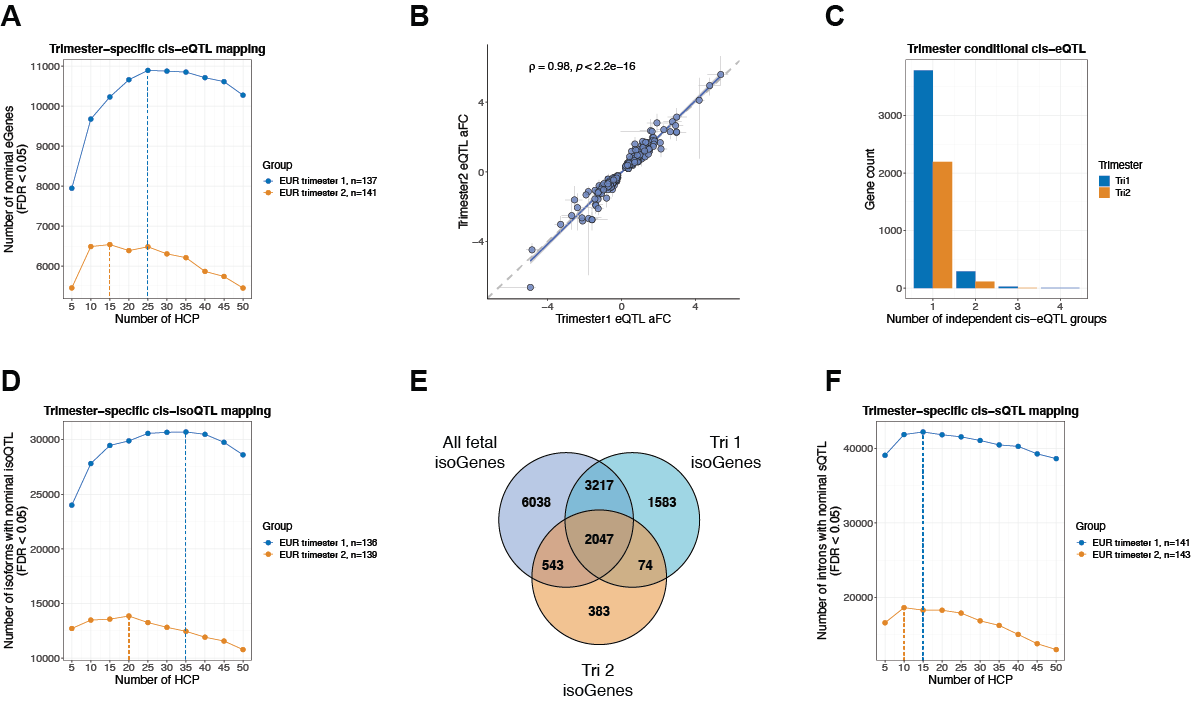

##### Fig. S7: Trimester-specific xQTL mapping.

**(A)** Trimester-specific *cis*-eQTL nominal mapping. 25 and 15 HCPs maximized the number of nominal eGenes and were included in a permutation-based analysis for the first and second trimesters, respectively. Note: trimester sample sizes differ slightly for e/iso/sQTL as gene and isoform expression outliers were removed from e/isoQTL mapping. In sQTL mapping, only genotype-related samples were removed. **(B)** Correlation of eQTL effect sizes between the two trimesters, as measured by allelic fold change (aFC) between the two trimesters. **(C)** Trimester 1 and 2 conditional eQTL mapping. **(D)** Trimester-specific *cis*-isoQTL nominal mapping. 20 and 35 HCPs maximized the number of isoforms with nominal isoQTLs and were included in a permutation-based analysis for the first and second trimesters, respectively. (**E**) Comparison of isoGenes discovered in Tri1, Tri2, and the full dataset. **(F)** Trimester-specific *cis*-sQTL nominal mapping. 15 and 10 HCPs maximized the number of introns with nominal sQTLs and were included in a permutation-based analysis for the first and second trimesters, respectively.

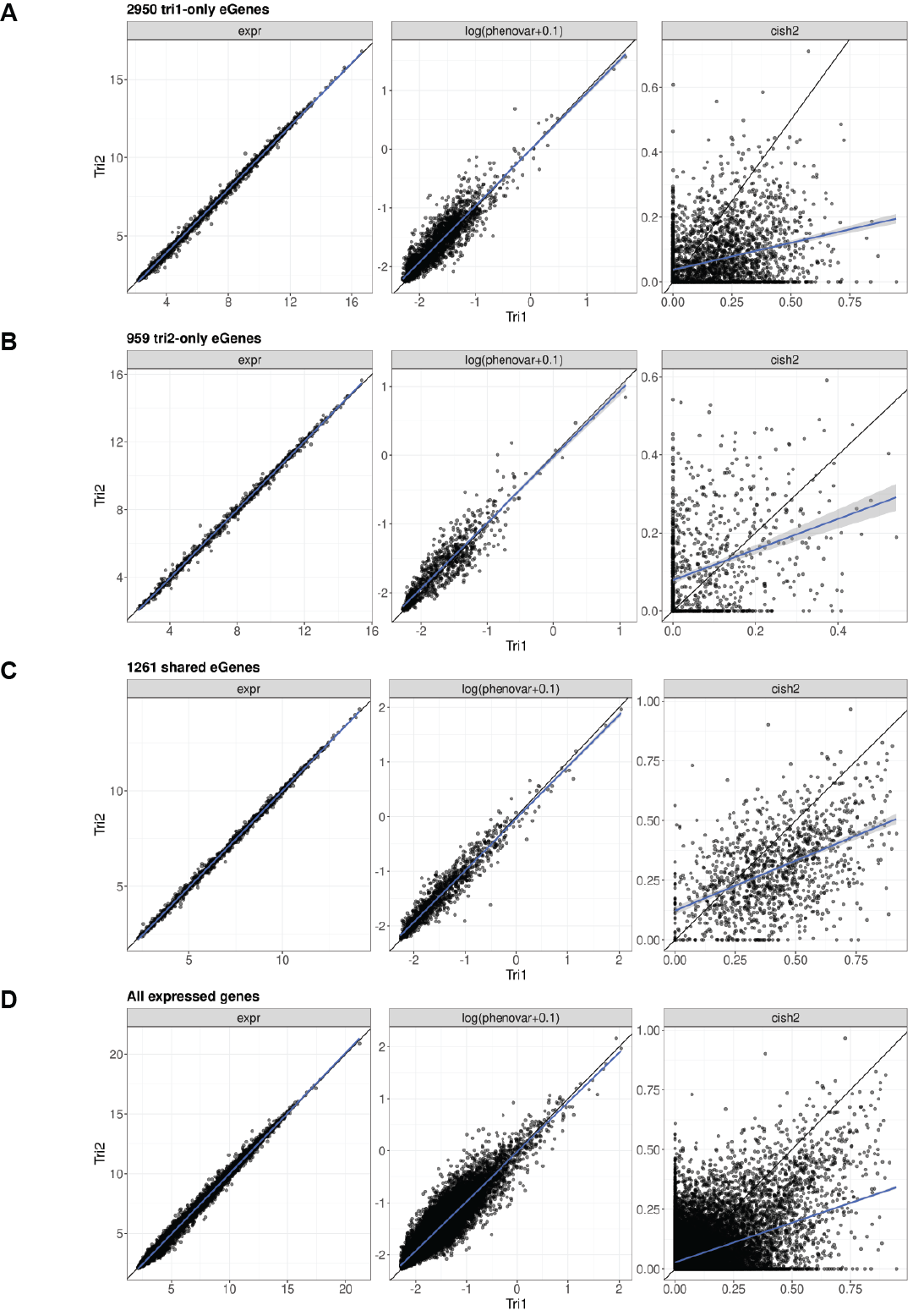

##### Fig. S8: Trimester heritability analysis of (A) trimester 1-only eGenes, (B) trimester 2-only eGenes, (C) trimester shared eGenes, and (D) all expressed genes. Gene expression level, phenotypic variance, and cish2 estimates in trimester 1 and 2 are shown on the x and y axes, respectively.

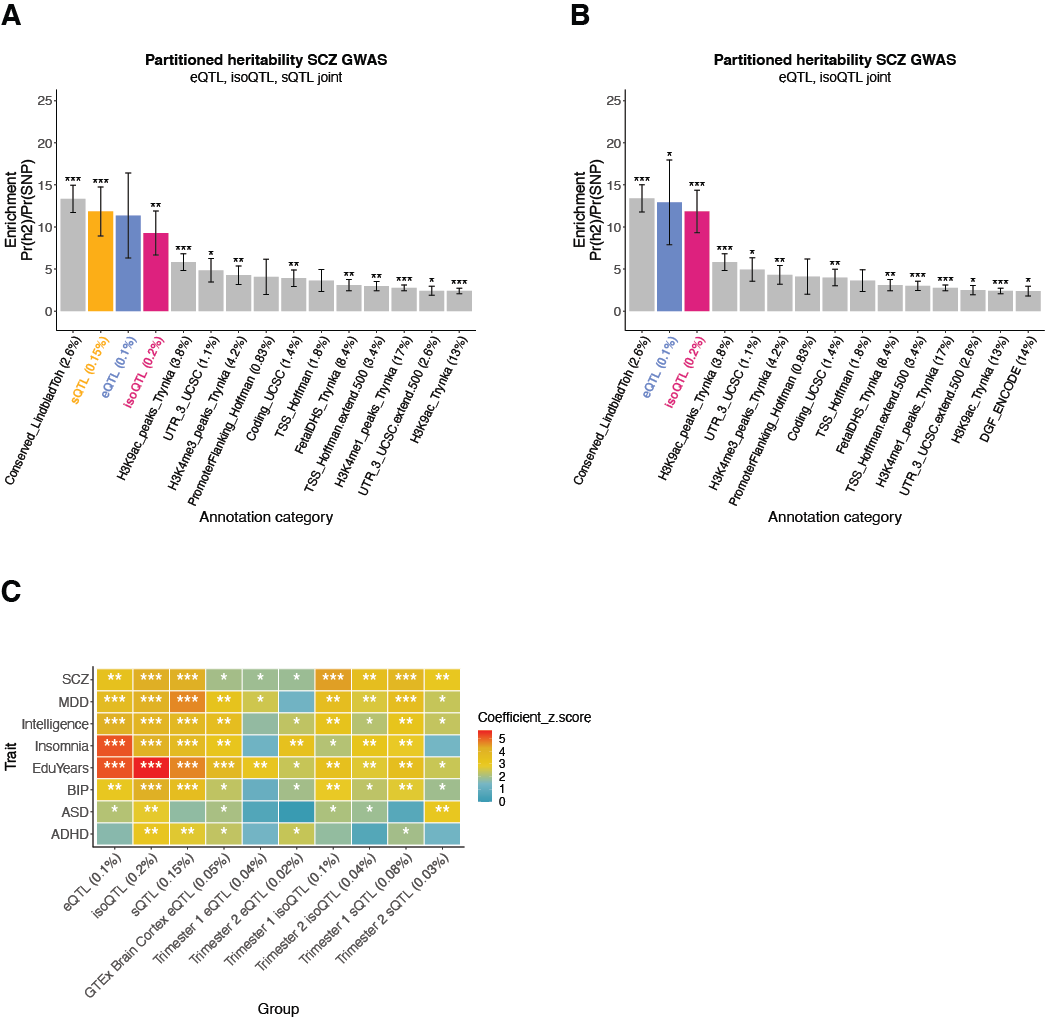

##### Fig. S9: S-LDSC

**(A)** S-LDSC enrichment of SCZ GWAS risk in fetal brain eQTL, isoQTL, sQTL, and background functional annotations, estimated jointly. For A, B, C: *** FDR<0.001, ** FDR<0.01, * FDR<0.05. In parentheses are the proportion of SNP of each annotation. **(B)** S-LDSC enrichment of SCZ GWAS risk in eQTL, isoQTL, and background annotations, estimated jointly. **(C)** S-LDSC coefficient z-scores of GWAS traits in various groups of brain QTLs.

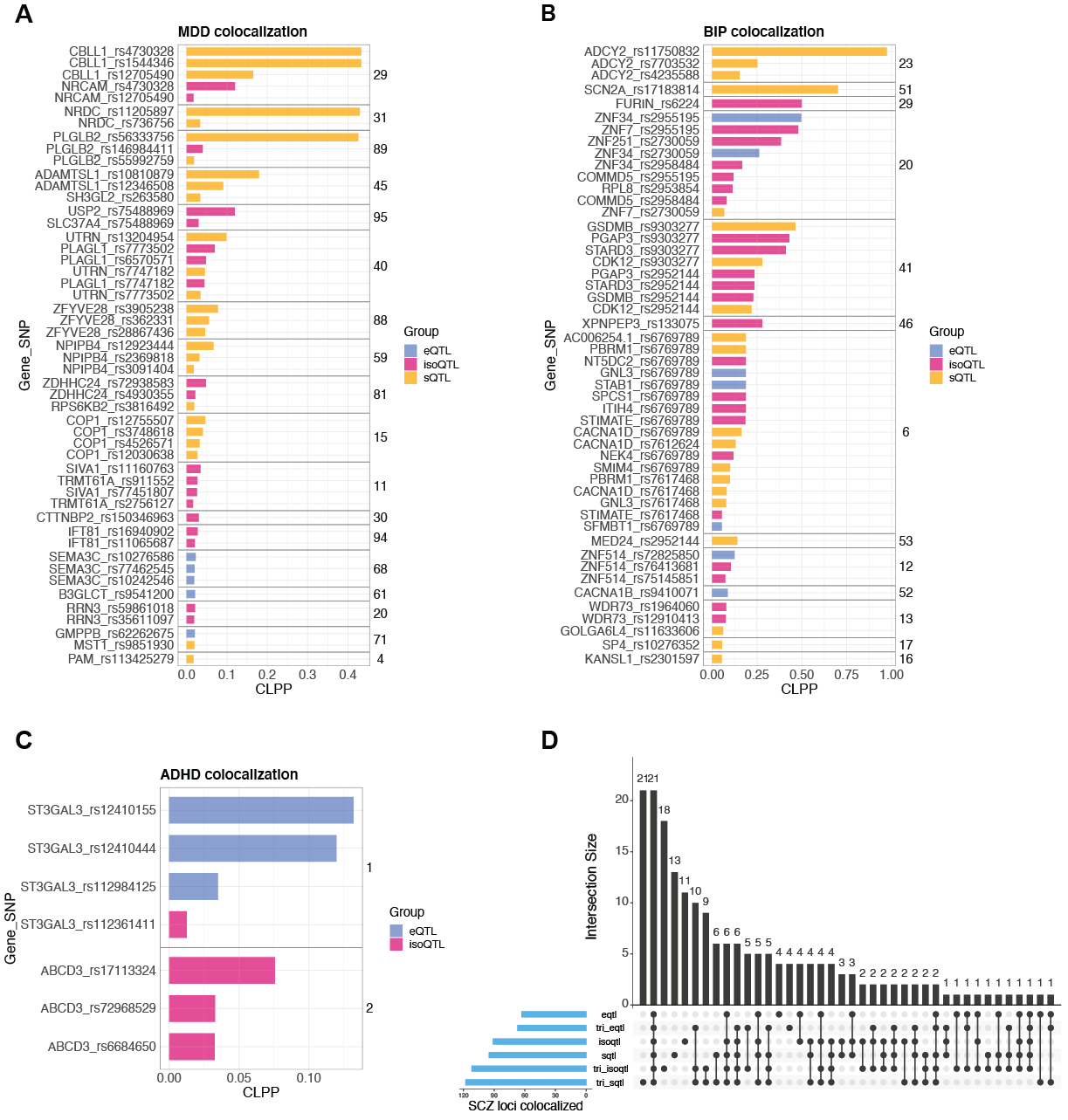

##### Fig. S10: Colocalization.

Top CLPP results for (**A**) MDD, (**B**) BIP, and (**C**) ADHD. Results are colored by QTL annotation. Numbers on the right represent GWAS loci. (**D**) Overlap of SCZ loci colocalized with bulk and trimester-specific e/iso/sQTL.

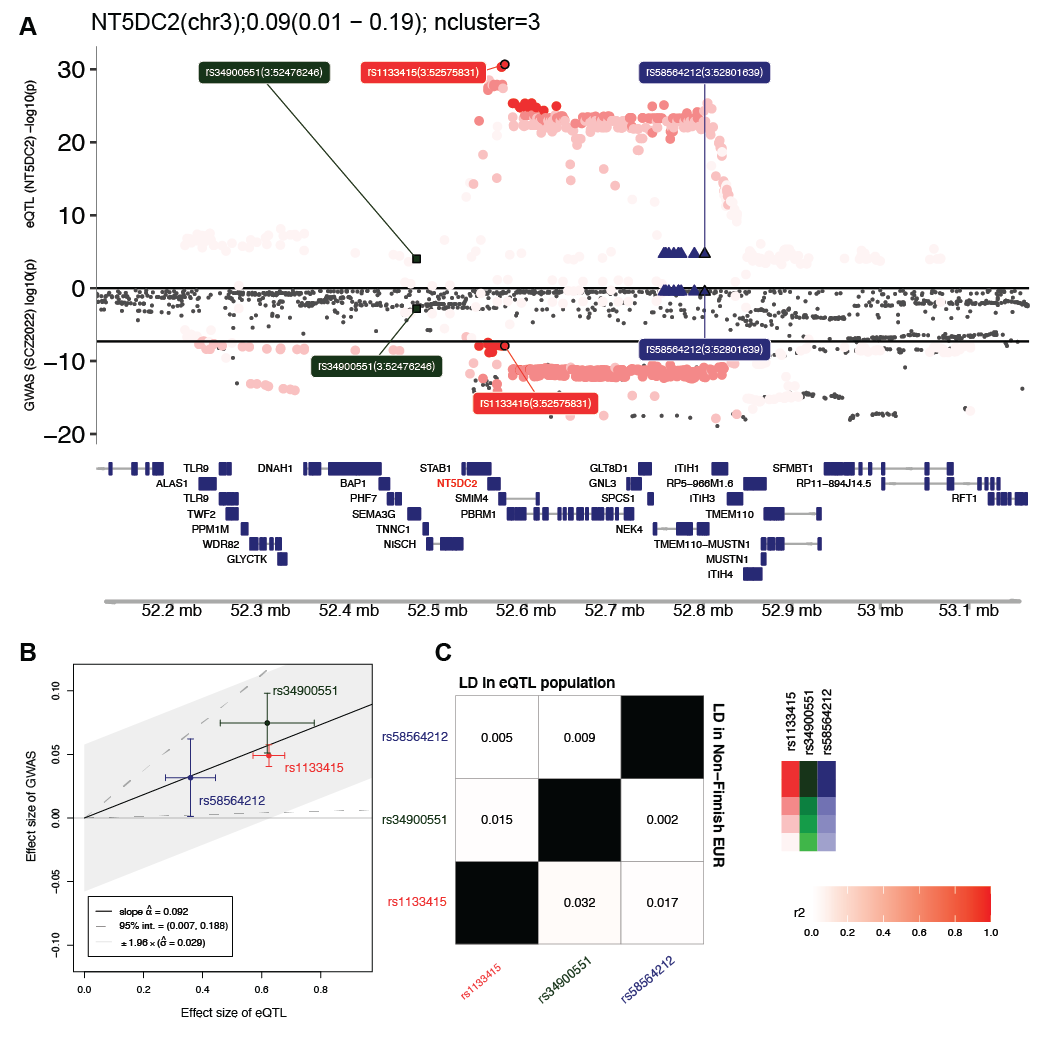

##### Fig. S11: MRLocus for NT5DC2 locus.

(**A**) NT5DC2 eQTL and SCZ GWAS associations are shown with a LocusZoom plot. Variants are colored by the 3 conditionally independent eQTL groups. (**B**) eQTL and GWAS effect sizes of the eCAVIAR-selected variants in the 3 eQTL groups. Bars represent the standard error of eQTL and GWAS effect sizes. (**C**) LD between the 3 eCAVIAR-selected variants, calculated in the multi-ancestry fetal brain dataset (left top) or non-Finnish EUR (right bottom), showing that these eQTL variants are nearly independent.

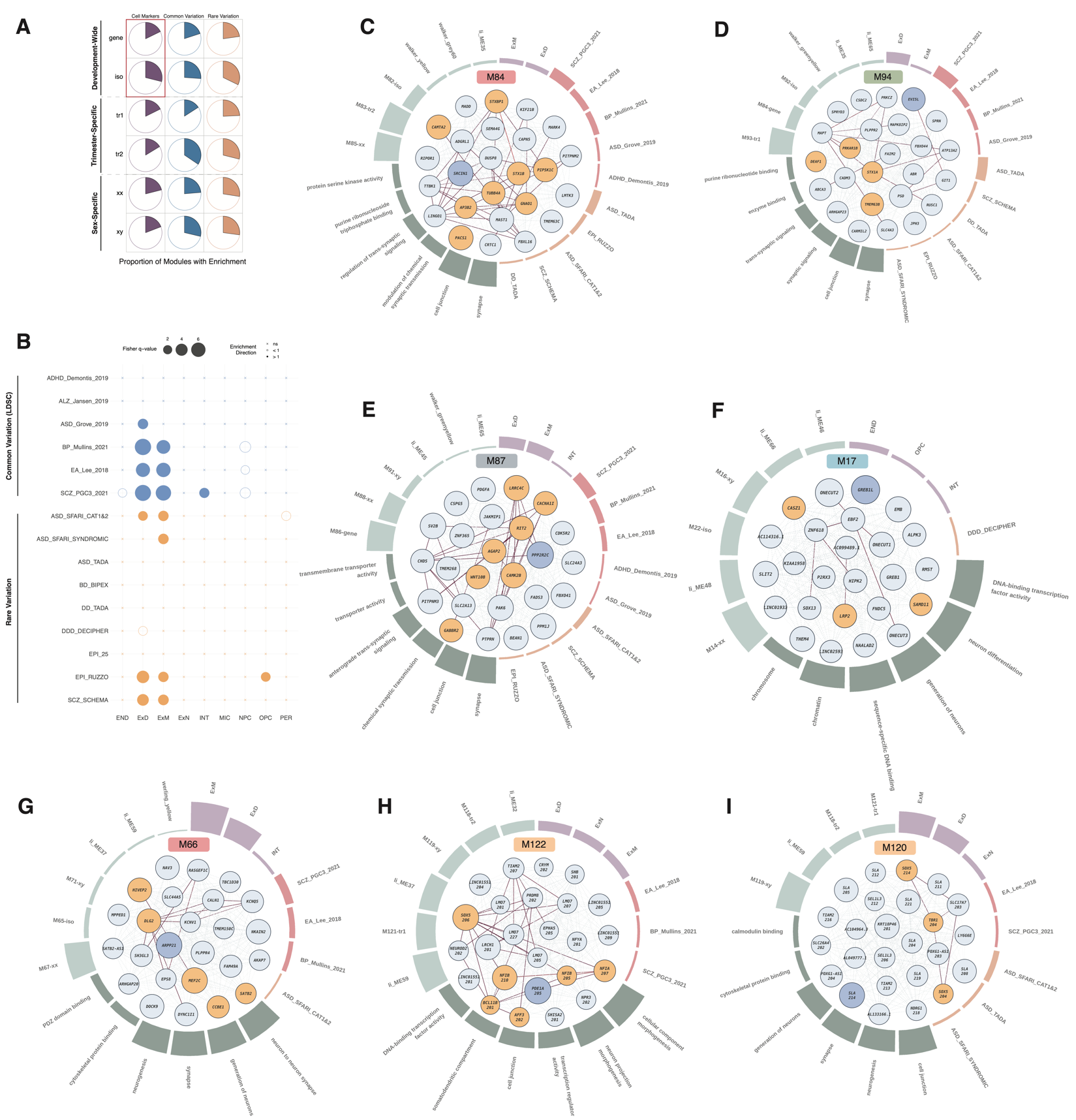

##### Fig. S12: Gene and isoform-level co-expression modules identified by WGCNA

(**A**) Proportion of modules exhibiting cell marker enrichment (purple), common variation enrichment (blue), and rare variation enrichment (orange), wherein red indicates statistically significant shifts in proportions **(B)** Association of rare variant-enriched modules with cell type markers (**C**) Hairball and barplot for M84, a development-wide gene module. Center: hairball of top 25 module genes by kME, dark blue indicates the module hub gene, orange indicates genes with associated high-risk rare variants. Solid edges indicate the level of experimental evidence for protein-protein interaction from the STRING database: 0 (purple) to 999 (red). Surrounding: circular bar plot highlighting module enrichment for cell types (purple), common (red) and rare (orange) variation, gene ontology terms (dark green), and module overlap (light green). (**D**) Hairball and barplot for M94, an XY module. (**E**) Hairball and barplot for M87, a trimester 2 module. (**F**) Hairball and barplot for M17, a trimester 1 module. (**G**) Hairball and barplot for M66, a gene module. (**H**) Hairball and barplot for M122, an isoform module. (**I**) Hairball and barplot for M120, an isoform module.

### Supplementary Tables

#### Table S1. Metadata and xQTL summary statistics

- README
- ST1-1-metadata: Metadata of 654 distinct, high-quality samples
- ST1-2-eGene: Sum stats of 10094 permutation eGenes and their primary *cis*-eQTL
- ST1-3-isoGene: Sum stats of 11845 group permutation isoGenes and their primary *cis*-isoQTL
- ST1-4-sGene: Sum stats of 7490 group permutation sGenes and their primary *cis*-sQTL
- ST1-5-torus-funcEnrich: Torus functional enrichment of *cis*-e/iso/sQTL
- ST1-6-conditional: Sum stats of conditional *cis*-eQTL mapping, for detailed descriptions of columns, see <https://qtltools.github.io/qtltools/>
- ST1-7-fetalOnlyeGene-CellTypeEnrich: Cell type enrichment for 2488 fetal only eGenes. "apprach" column denotes whether the cell types are defined as broad, cluster, or sub-cluster
- ST1-8-inv-eQTL: Sum stats of significant inversion eQTL

#### Table S2. Population specific xQTL and fine-mapping

- README
- ST2-1-EUR-eGenes: Sum stats of 5296 EUR permutation eGenes and their primary *cis*-eQTL
- ST2-2-AMR-eGenes: Sum stats of 3242 EUR permutation eGenes and their primary *cis*-eQTL
- ST2-3-AFR-eGenes: Sum stats of 1876 EUR permutation eGenes and their primary *cis*-eQTL
- ST2-4-EUR-isoGenes	: Sum stats of 11672/6885 isoforms/isoGenes with permutation *cis*-isoQTL in EUR
- ST2-5-AMR-isoGenes: Sum stats of 5747/3989 isoforms/isoGenes with permutation *cis*-isoQTL in AMR
- ST2-6-AFR-isoGenes: Sum stats of 3719/2682 isoforms/isoGenes with permutation *cis*-isoQTL in AFR
- ST2-7-EUR-sGenes: Sum stats of 16277/4963 introns/sGenes with permutation *cis*-sQTL in EUR
- ST2-8-AMR-sGenes: Sum stats of 6324/2772 introns/sGenes with permutation *cis*-sQTL in AMR
- ST2-9-AFR-sGenes: Sum stats of 4337/2069 introns/sGenes with permutation *cis*-sQTL in AFR

#### Table S3. Trimester xQTL summary statistics

- README
- ST3-1-tri1-eGene: Sum stats of 4211 permutation eGenes in Tri1
- ST3-2-tri2-eGene: Sum stats of 2220 permutation eGenes in Tri2
- ST3-3-tri1-isoGene: Sum stats of 10881/6921 isoforms/isoGenes with permutation *cis*-isoQTL in Tri1
- ST3-4-tri2-isoGene: Sum stats of 4179/3047 isoforms/isoGenes with permutation *cis*-isoQTL in Tri2
- ST3-5-tri1-sGene: Sum stats of 14193/5312 introns/sGenes with permutation *cis*-sQTL in Tri1
- ST3-6-tri2-sGene: Sum stats of 5348/2318 introns/sGenes with permutation *cis*-sQTL in Tri2
- ST3-7-triOnly-biotype: Counts of gene types of tri1 and tri2-only e/sGenes
- ST3-8-triOnly-cellTypeEnrich: Cell type enrichment of tri1 and tri2-only e/sGenes

#### Table S4. S-LDSC and MESC results

- README
- ST4-1-sLDSC: S-LDSC of background and various brain QTL annotations (e/iso/sQTL, GTEx brain cortex eQTL, tri e/iso/sQTL)
- ST4-2-sLDSC-e/iso/sQTL-joint: S-LDSC jointly running background annotations, and e+iso+sQTL, top enrichment 15 annotations
- ST4-3-sLDSC-e/isoQTL-joint: S-LDSC jointly running background annotations, and e+isoQTL, top enrichment 15 annotations
- ST4-4-MESC: h2med results for all expressed genes/isoforms/introns in the full dataset, and trimester specific datasets

#### Table S5. isoTWAS and colocalization

- README
- ST5-1a-isoTWAS-preFOCUS: isoTWAS statistics of isoforms that are permutation significant
- ST5-1b-isoTWAS-finemapped: isoTWAS statistics of isoforms that are in 90% FOCUS credible sets
- ST5-2 to ST5-29: eCAVIAR significant colocalization results for various neuropsychiatric GWAS and fetal brain QTLs
- ST5-30: MRLOCUS summary statistics

#### Table S6. Network analyses

- README
- ST6-1-Module-Genes: Membership of genes and transcripts to each of the modules
- ST6-2-All-Enrichment: Compiled module enrichment data from ST6-3, ST6-4, and ST6-5
- ST6-3-Cell-Enrichment: Cell type enrichment of modules using Polioudakis fetal brain marker genes
- ST6-4-Common-Enrichment: Common variation enrichment of modules using sLDSC and MAGMA
- ST6-5-Rare-Enrichment: Rare variation enrichment of modules using logistic regression
- ST6-6-Module-Overlap: Overlap of fetal modules with other fetal modules in the dataset or modules from Walker et al. (walker_), Werling et al. (werling_), and Li et al. (li_)

#### Table S7. CellWalker and module interacting eQTL summary statistics

- README
- ST7-1-CellWalker-single-label: CellWalker mapping of bulk eQTL SuSiE results to single cell type labels. Corresponds to Fig 7A
- ST7-2-CellWalker-ML-strict: CellWalker mapping of bulk eQTL SuSiE results to multi-level ("ML") cell type labels. Filtered for number of cell types. See methods for details

### Supplementary Data

Data S1 (Mixed_ciseqtl_90hcp_perm_purity_filtered.txt.gz): SuSiE fine-mapping results for multi-ancestry *cis*-eQTL

Data S2 (Eur_ciseqtl_50hcp_perm_purity_filtered.txt.gz): SuSiE fine-mapping results for EUR *cis*-eQTL

Data S3 (Amr_ciseqtl_15hcp_perm_purity_filtered.txt.gz): SuSiE fine-mapping results for AMR *cis*-eQTL

Data S4 (Afr_ciseqtl_25hcp_perm_purity_filtered.txt.gz): SuSiE fine-mapping results for AFR *cis*-eQTL

Data S5 (EUR_tri1_gene_cish2.csv): heritability estimates for genes in EUR Tri1 samples

Data S6: EUR_tri2_gene_cish2.csv): heritability estimates for genes in EUR Tri2 samples

Data S7: (EUR_tri1_splicing_cish2.csv): heritability estimates for local splicing in EUR Tri1 samples

Data S8: (EUR_tri2_splicing_cish2.csv): heritability estimates for local splicing in EUR Tri2 samples

Data S9 (modeqtl_top.RData): module interaction eQTL permutation eGenes and their primary ieQTLs
